# Cross-ancestry genome-wide association study and systems-level integrative analyses implicate new risk genes and therapeutic targets for depression

**DOI:** 10.1101/2023.02.24.23286411

**Authors:** Yifan Li, Xinglun Dang, Rui Chen, Junyang Wang, Shiwu Li, Brittany L. Mitchell, Yong-Gang Yao, Ming Li, Tao Li, Zhijun Zhang, Xiong-Jian Luo

## Abstract

Deciphering the genetic architecture of depression is pivotal for characterizing the associated pathophysiological processes and development of new therapeutics. Here we conducted a cross-ancestry genome-wide meta-analysis on depression (416,437 cases and 1,308,758 controls) and identified 287 risk loci, of which 140 are new. Variant-level fine-mapping prioritized potential causal variants and functional genomic analysis identified variants that regulate the binding of transcription factors. We validated that 80% of the identified functional variants are regulatory variants and expression quantitative trait loci (eQTL) analysis uncovered the potential target genes regulated by the prioritized risk variants. Gene-level analysis, including transcriptome-wide association study (TWAS), proteome-wide association study (PWAS), colocalization and Mendelian randomization-based analyses, prioritized potential causal genes and drug targets. Combining evidence from different analyses revealed likely causal genes, including *TMEM106B, CTNND1, EPHB2, AREL1, CSE1L, RAB27B, SATU1, TMEM258, DCC, etc*. Pathway analysis showed significant enrichment of depression risk genes in synapse-related pathways. Finally, we showed that *Tmem106b* knockdown resulted in depression-like behaviors in mice, supporting involvement of *Tmem106b* in depression. Our study identified new risk loci, likely causal variants and genes for depression, providing important insights into the genetic architecture of depression and potential therapeutic targets.

## Introduction

Depression is one of the most prevalent mental disorders^1^ and a leading cause of disability worldwide^2^. Though recent genome-wide association studies (GWASs) have reported multiple risk loci for depression^3-8^, much of the underlying heritability remains unexplained. In addition, most depression GWASs have been conducted in populations of European ancestry, potentially missing important genetic insights into depression. More importantly, the causal variants and genes remain largely unknown for most reported risk loci, hampering the translation of genetic findings into clinical applications and therapeutics. Hence, the discovery of new genetic risk loci and functional characterization of the identified risk variants and genes will provide important insights into depression pathophysiology and therapeutic targets.

In this study, we first conducted a large-scale cross-ancestry meta-analysis (416,437 cases and 1,308,758 controls) on depression. Based on the results of the meta-analysis, we conducted comprehensive prioritization and integrative analysis to prioritize the potential causal variants and genes. We prioritized potential causal variants and we validated the regulatory effect of the identified functional variants. In addition, we also prioritized likely causal genes, including *TMEM106B, CTNND1, EPHB2, etc*. Finally, we found that *Tmem106b* knockdown resulted in depression-like behaviors in mice, providing animal model evidence that supports *TMEM106B* is a depression risk gene. Our study identified new risk loci, likely causal variants, and genes for depression, providing important insights into the genetic architecture of depression and potential therapeutic targets.

## Results

### Cross-ancestry genome-wide meta-analysis identified 287 risk loci for depression

We conducted a meta-analysis by combining genome-wide associations reported by 6 previous studies (**Methods**), including Million Veteran Project (MVP) (two cohorts were included, cohort of European ancestry, 83,810 cases and 166,405 controls; cohort of African ancestry, 25,843 cases, and 33,757 controls)^3^, FinnGen (https://www.finngen.fi/en) (33,812 cases and 271,380 controls), studies reported by Howard *et al*. (UKB+PGC+23andme) (246,363 cases and 561,190 controls)^5^, the Australian Genetics of Depression Study (AGDS) (13,318 cases and 12,684 controls)^9^, Giannakopoulou *et al*. (12,455 cases and 85,548 controls)^10^, and Sakaue *et al*. (Biobank Japan, 836 cases and 177,794 controls)^11^. Meta-analysis (a total of 416,437 cases and 1,308,758 controls) identified 287 independent genomic risk loci (*P* < 5×10^−8^), of which 140 were new (genome-wide significant SNPs that have not been reported by each included study and previous depression GWASs^3-5,9-11^were considered as new associations.) (**Fig.1, Supplementary Table S1** and **S2**). The most significant SNP rs7531118 is located approximately 89 kb downstream of the *NEGR1* (1p31.1) (**Fig.2a**), and the second most significant SNP rs1021363 is located in the intron 2 of the *SORCS3* (**Fig.2b**). Linkage disequilibrium score regression (LDSC) showed that polygenicity rather than confounding factors account for most of the associations (lambda GC = 1.54, intercept (s.e.) = 1.04 (0.01), Ratio (s.e.) = 0.03 (0.008)). These findings expand the risk loci of depression substantially.

**Fig. 1.**
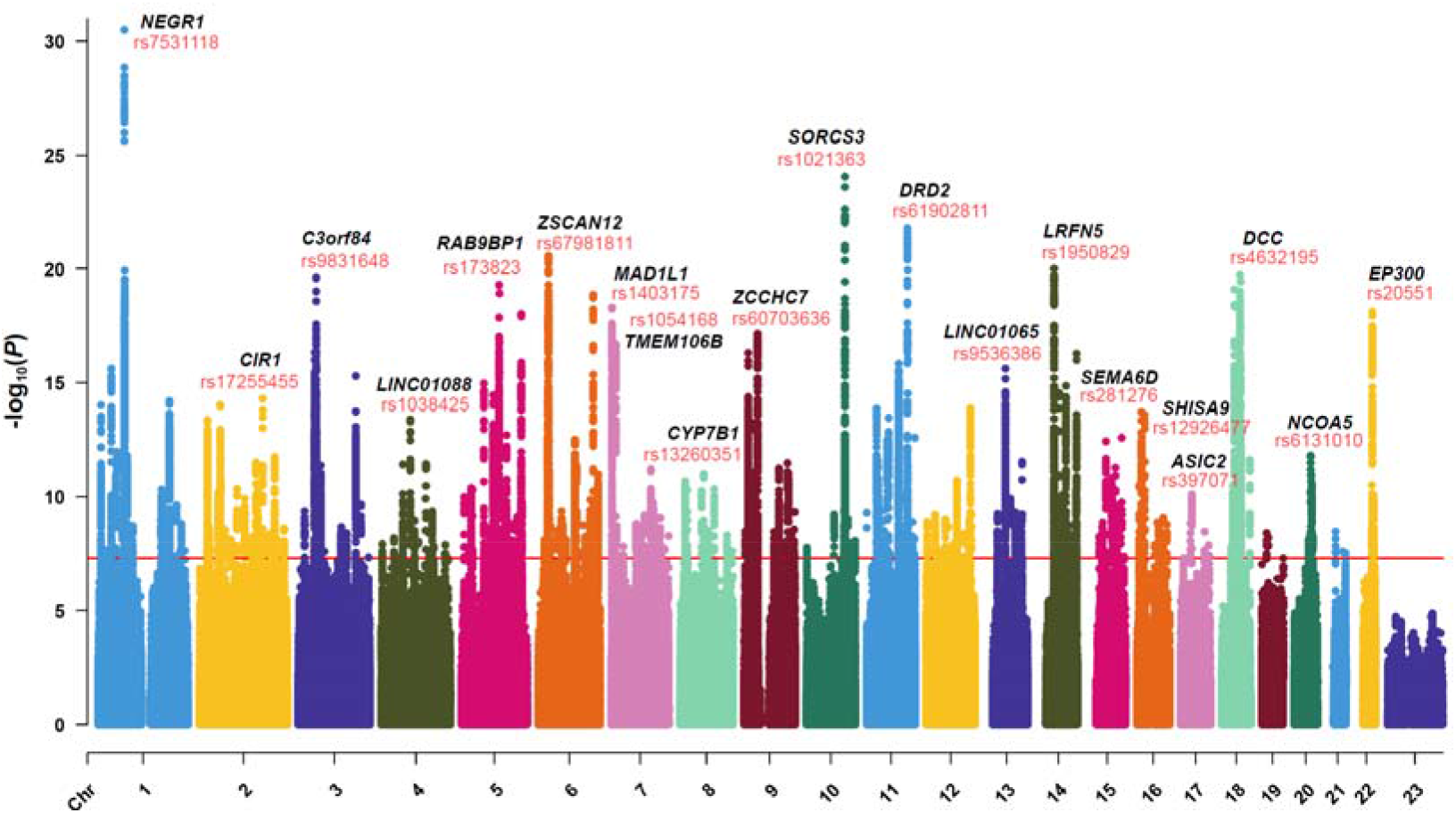
Manhattan plot of the GWAS meta-analysis. Associations of the GWAS meta-analysis (N =416,437 cases and 1,308,758 controls). The red line shows the genome-wide significant *P* threshold (*P* < 5.0 × 10^−8^). SNP rs7531118 near *NEGR1* showed the most significant association. Only the top 20 risk loci are shown. The lead SNPs are shown in red, and the nearest genes to the lead SNPs are shown in bold italic text.

**Fig. 2.**
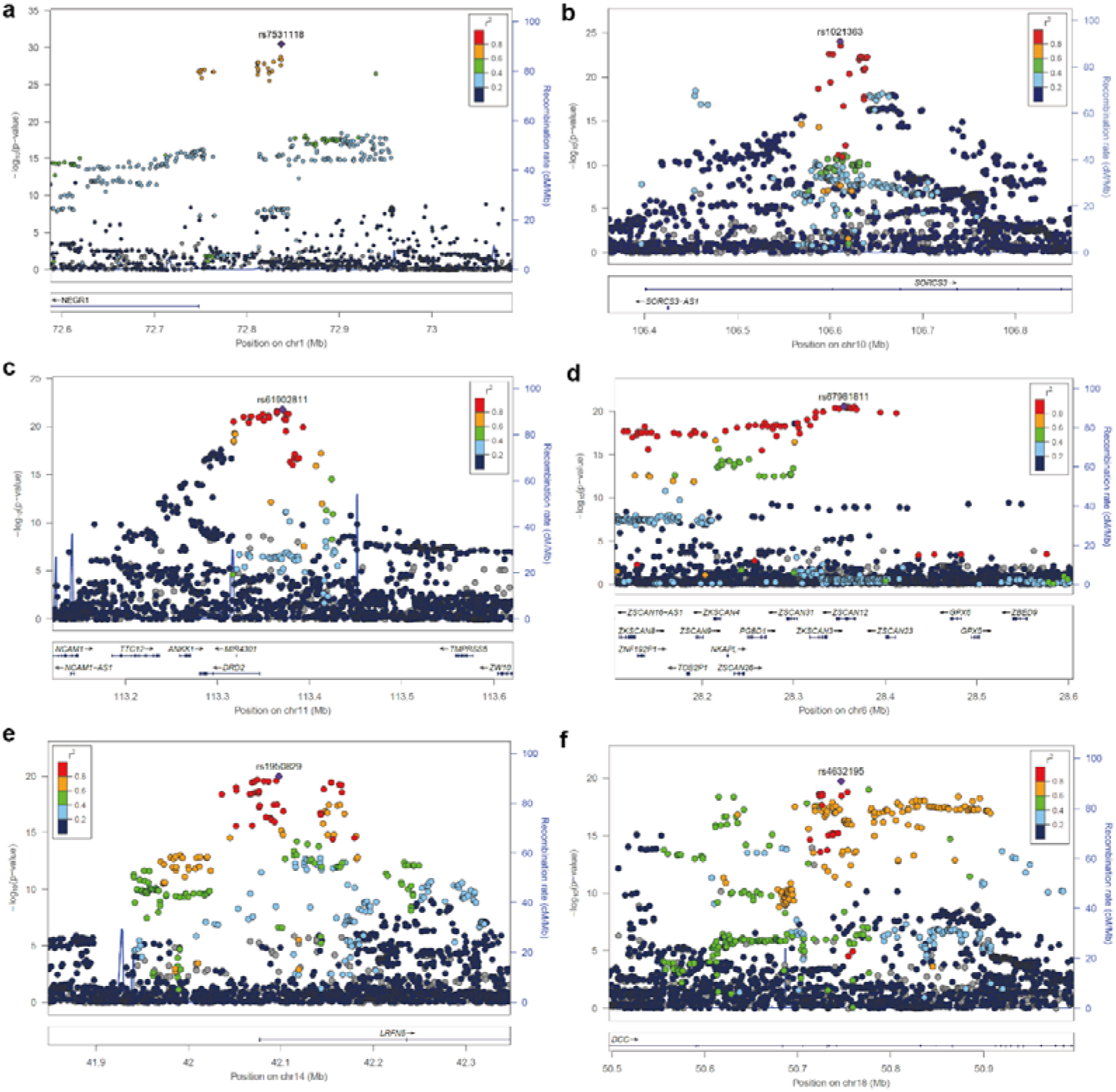
Locuszoom plots for six genome-wide significant association loci. rs7531118 is located in downstream of *NEGR1*, rs1021363 is located in intronic of *SORCS3*, rs61902811 is located in downstream of *DRD2*, rs67981811 is located in UTR3 of *ZSCAN12*, rs1950829 is located in intronic region of *LRFN5*, and rs4632195 is located in intronic of *DCC*.

### Heritability and genetic correlations

SNP heritability (h^2^) estimate based on our meta-analysis results was 0.034 (s.e. = 0.003) (estimated by linkage disequilibrium score regression, LDSC^12^) and 0.043 (s.e. = 0.002) (calculated by SumHer^13^ from the LDAK software package, with the use of the BLD-LDAK model). This value is slightly lower than previously reported, as expected^5,9^. We calculated the genetic correlations between the six included depression studies (MVP, FINNGEN, AGDS, 23me_UKB_PGC, Howard *et al*., and Sakaue *et al*.), and found that the genetic correlations between different depression studies are highly significant, except for two studies by Giannakopoulou *et al*.^10^ and Sakaue *et al*.^11^ (**Supplementary Table S3)**. For these two studies, the SNP heritability is very low (0.0104 and -0.0014, respectively). Of note, we failed to calculate meaningful genetic correlations between the study by Sakaue *et al*.^11^ and other studies. A possible reason for this is the small number of cases included in this study (only 836 cases). The genetic correlations were higher between cohorts of European ancestry, whereas the genetic correlations were much lower between cohorts of African and Asian ancestry, suggesting differences in genetic background between different datasets. However, evidence based on LD intercepts (1.04, s.e. = 0.010) and attenuation ratio (0.03, s.e. = 0.008) showed negligible inflation or confounding in our meta-analysis results (**Supplementary Table S4**). Finally, we investigated the genetic correlations between depression (i.e., the combined meta-analysis results of the six different depression studies) and other brain disorders and intelligence (**Supplementary Table S5**). The top three disorders that showed the most significant genetic correlations with depression were anxiety disorders, post-traumatic stress disorder (PTSD), and neuroticism (**Supplementary Fig.S3**). Most of the analyzed traits showed positive correlations with depression. However, intelligence and Parkinson’s disease showed negative correlations.

### Functional genomics identified potential causal variants at the risk loci

Identifying functional (or causal) variants is crucial for follow-up mechanistic investigation and functional characterization. To identify the functional (or potential causal) variants from the identified risk loci, we conducted a functional genomic analysis, as previously described^14-16^. We identified 64 functional SNPs that affect the binding of transcription factors (i.e., TF binding-affecting SNPs) (**Supplementary Table S6**). Among these 64 TF binding-affecting SNPs, 37 affected the binding of CTCF and 10 affected the binding of REST, and 6 SNPs interfered with more than one TFs simultaneously (**Fig.3, Supplementary Table S6**). About 40% TF binding-affecting SNPs are located in the intronic region (**Fig.3a**), indicating the pivotal role of intronic variants in depression. Notably, 34 TF-disrupting SNPs showed genome-wide significant (GWS) associations with depression, and the CTCF binding-affecting SNP rs7531118 (located downstream of the *NEGR1*) showed the most significant association (**Supplementary Table S6**). These results pinpointed the functional (or potential causal) variants from the reported risk loci and suggested that affecting the binding of TFs is a major manner by which these functional SNPs exert their biological effect on depression.

**Fig. 3.**
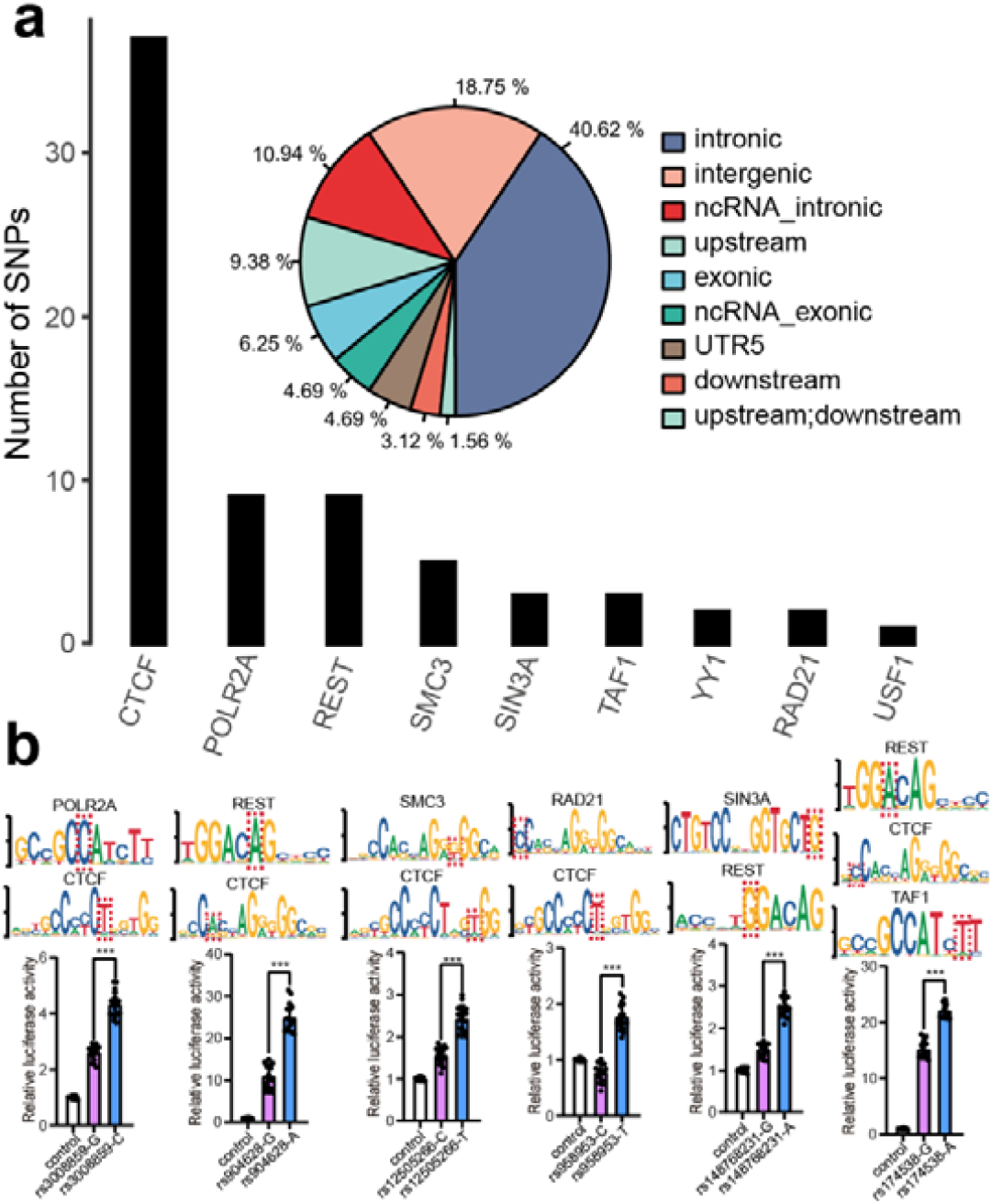
Single-nucleotide polymorphisms (SNPs) that affect binding of transcription factors. **a**, **Left panel**, The box plot shows the number of depression risk SNPs that affect the binding of individual TF based on matched position weight matrices (PWMs) from chromatin immunoprecipitation and sequencing (ChIP-Seq) and PWM database. **Right panel**, The distribution of the 64 identified TF binding–affecting SNPs in the human genome. A large proportion of the identified regulatory SNPs were located in intronic regions. **b, SNPs that affect the binding of two or three transcription factors. Upper panel**, the position weight matrix (i.e., binding motif) of the corresponding TFs. **Lower panel**, results of reporter gene assays. The Renilla internal control was used to normalize the luciferase activity, and the *y* axis shows the relative luciferase activity. For each group, 16 replicates were used (n = 16). Two-tailed *Student’s t test* was used to compare if the difference reaches the significance level (*P* < 0.05). Data represent mean sd. The red dashed box highlights the test SNP. Different alleles of all SNPs showed significant differences in luciferase activity, indicating these SNPs are functional variants.

### Reporter gene assays validated that 80% of the TF binding-affecting SNPs are regulatory variants

To validate the regulatory effect of the identified TF binding-affecting SNPs, we conducted dual-luciferase reporter gene assays for all TF binding-affecting SNPs (**Supplementary Table S6**). Reporter gene assays revealed that 50 out of 63 SNPs (vector for one SNP were not successfully constructed due to complex genomic sequence) showed regulatory effect, i.e., different alleles of these 50 SNPs affected the luciferase activity significantly (*P* < 0.05) (**Fig.4, Supplementary Fig.S8**). These results provided experimental evidence that supports most of the identified TF binding-affecting SNPs are regulatory variants. Considering the interaction between TFs and regulatory sequence has a crucial role in expression regulation, these findings also indicate that these functional SNPs may confer risk of depression by regulating gene expression. These results provide important insights into the regulatory mechanisms of depression risk variants.

**Fig. 4.**
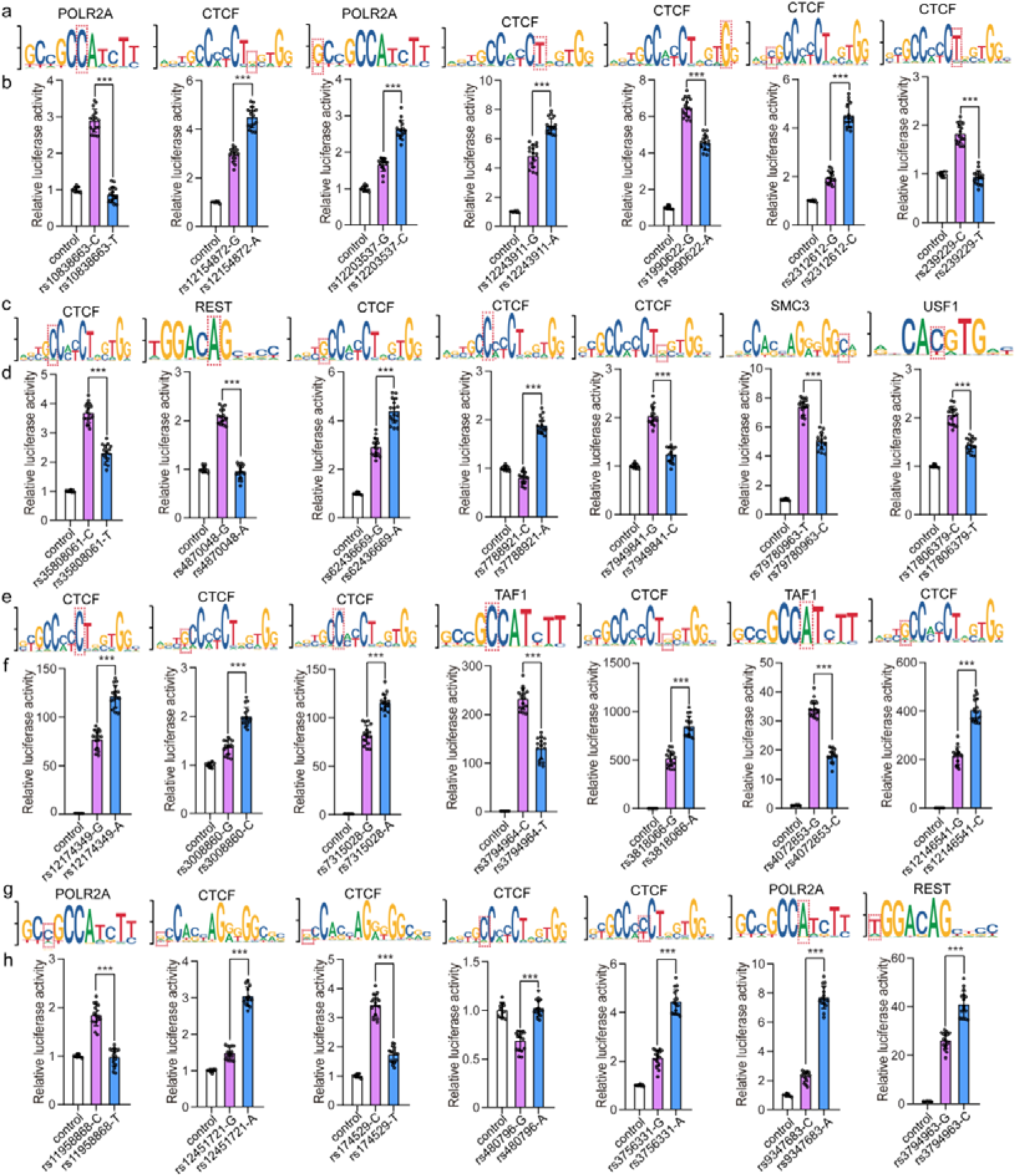
Reporter gene assays validated the regulatory effect of the identified TF binding-affecting SNPs. The upper panel shows the binding motifs (position weight matrix, PWM) of the corresponding TFs. The lower panel shows the results of reporter gene assays. DNA fragments (about 600 bp) containing different alleles of the TF binding-affecting SNPs were amplified and cloned into the pGL4.11-basic vector (for promoter activity detection) or pGL3-promoter vector (for enhancer activity detection). For each pair of constructs, there was only one nucleotide difference at the test SNP, so the reporter gene assays will reveal if different alleles of the test SNP cause differences in luciferase activity. The Renilla internal control was used to normalize the luciferase activity, and the *y* axis shows the relative luciferase activity. For each group, 16 replicates were used (n = 16). Two-tailed *Student’s t test* was used to compare if the difference reaches the significance level (*P* < 0.05). Data represent mean sd. The red dashed box highlights the test SNP.

### Fine-mapping prioritized potential causal variants

To identify the potential causal variants from the identified risk loci, we utilized two well-characterized fine-mapping approaches, FINEMAP^17^ and SuSiE^18,19^, for causal variant fine-mapping. FINEMAP (k=3, assuming each locus contains three causal variants) prioritized potential causal variants with a posterior probability greater than 0.7 for 210 risk loci (**Supplementary Table S10**). SuSiE also prioritized high-confidence potential causal variants (posterior inclusion probability (PIP) >0.7) for 243 risk loci (**Supplementary Table S11**). Notably, both FINEMAP and SuSiE prioritized 82 identical SNPs as potential causal variants (**Supplementary Table S12**), suggesting that these SNPs are potential causal SNPs.

### eQTL analysis identified potential target genes of the risk variants

Most of the identified depression risk variants are located in non-coding region, implying regulation of gene expression is a major manner that the risk variants exert their effect. To identify potential target genes of the identified risk variants, we used the BrainMeta v2^20^ dataset, which contains eQTL data from 2,865 human brain transcriptome, to identify associations between risk variants and gene expression in the human brain. Lead SNPs, TF binding-affecting SNPs identified by functional genomics, and credible causal SNPs prioritized by FINEMAP and SuSiE were used for eQTL analysis. For lead SNPs, 285 showed nominally significant associations (uncorrected *P* < 0.05) with gene expression in the human brain, and 161 showed significant associations when adjusted by Bonferroni correction (*P* < 3.90×10^−05^) (**Supplementary Table S13**). We next examined the associations between the TF binding-affecting SNPs (**Supplementary Table S6**) and gene expression. Among the 64 TF binding-affecting SNPs, 50 showed significant associations with gene expression in the human brain (Bonferroni corrected *P* < 2.40×10^−05^) (**Supplementary Table S14**). Finally, we found that 82 SNPs prioritized by both FINAMAP and SuSiE showed significant associations with gene expression in the human brain (**Supplementary Table S12**). Taken together, these results identified the potential target genes regulated by the lead and prioritized functional risk SNPs, suggesting that these functional variants confer the risk of depression through regulating expression of these target genes.

### TWAS identified genes whose genetically regulated expression are associated with depression

By integrating the association results of the meta-analysis and brain eQTL data from the PsychENCODE^21^, we performed a TWAS (using FUSION^22^) to identify genes whose genetically regulated expression levels are associated with depression. TWAS identified 159 genes whose genetically regulated expression are associated with depression (Bonferroni corrected *P* < 2.86 × 10^−05^) (**Fig.5a**). Transcriptome-wide significant (TWS) genes include *RPL31P12, AREL1, TMEM106B, RHOA, RP11-73M18*.*6, etc*. Of note, *TMEM106B* (near the lead SNP rs1054168) showed the third significant association in TWAS. Besides, functional genomics also revealed that the functional SNP rs1990622 located downstream of *TMEM106B* showed strong association with depression (*P* = 2.30 × 10^−16^) (**Supplementary Table S6**). Consistent with functional genomic and eQTL analysis, reporter gene assays validated the regulatory effect of rs1990622, with G allele of rs1990622 conferred significantly higher luciferase activity compared with A allele (*P* = 7.56 × 10^−13^, **Fig.4**). These findings not only identified risk genes whose genetically regulated expression is associated with depression but also prioritized rs1990622 as a functional risk variant that confers risk of depression by regulating *TMEM106B* expression.

**Fig. 5.**
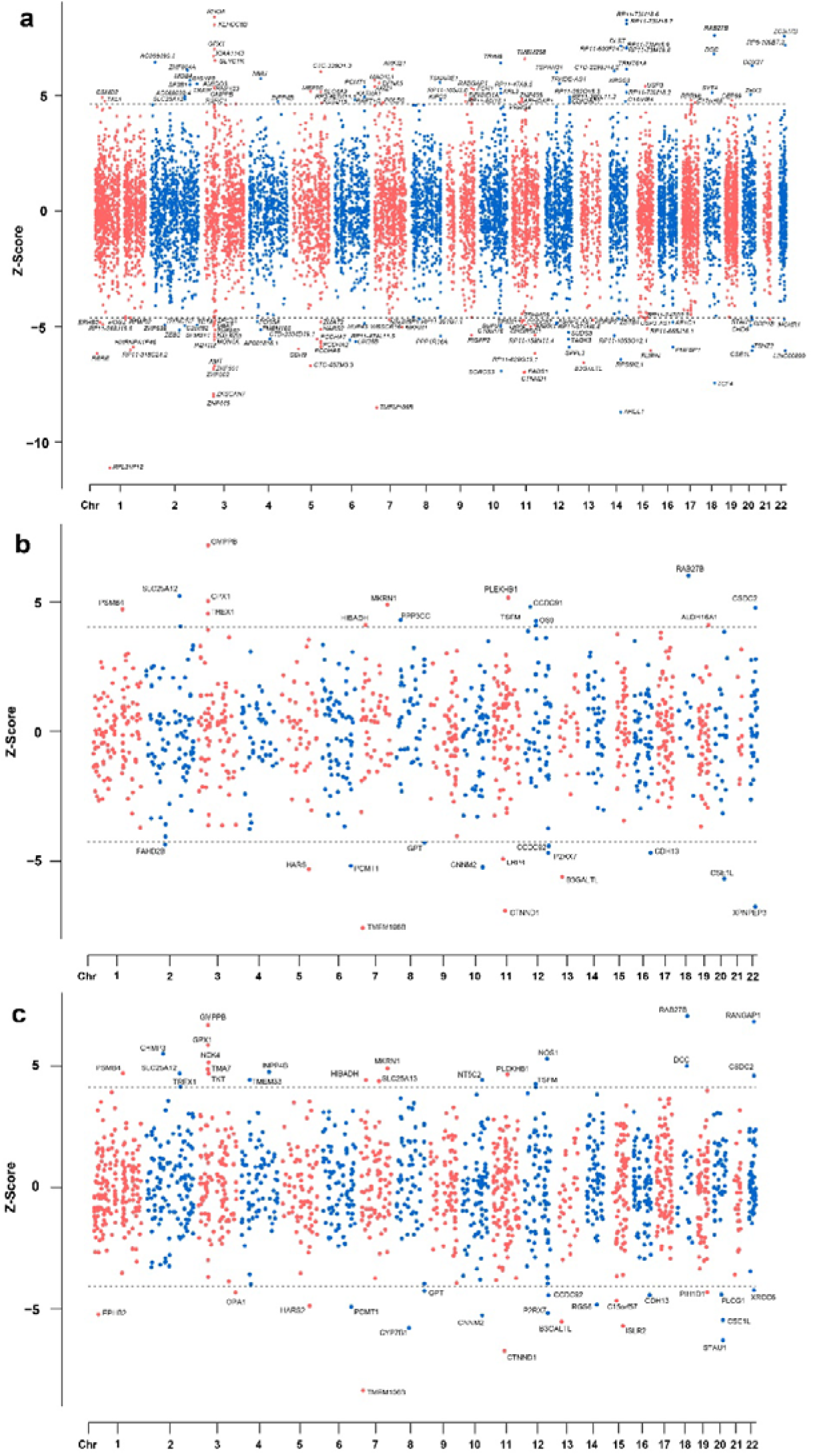
Transcriptome-wide and proteome-wide association results. Only transcriptome-wide and proteome-wide significant genes (Bonferroni corrected *P* < 0.05) are shown. **(a)** TWAS results. **(b**,**c)** PWAS results. Z > 0 indicates that elevated expression of this gene is associated with risk of depression, and Z < 0 indicates that lower expression of this gene is associated with risk of depression.

### PWAS nominated proteins whose genetically regulated abundance in the human brain are associated with depression

By integrating meta-analysis results and two independent human brain protein quantitative trait datasets (ROSMAP and Banner)^23^, we carried out PWASs. PWAS identified 51 proteins whose genetically regulated abundance were associated with depression (Bonferroni corrected *P* < 4.39 × 10^−5^ for Banner dataset, and *P* < 2.88 × 10^−5^ for ROSMAP dataset) (**Fig.5b, c**). Of note, 16 proteome-wide significant proteins also showed significant associations at transcriptome-wide level, including B3GALTL, CSE1L, CTNND1, DCC, EPHB2, GMPPB, GPX1, HARS, INPP4B, LRP4, MKRN1, PCMT1, RAB27B, SLC25A12, STAU1, and TMEM106B, strongly suggesting these genes are promising risk genes for depression. Interestingly, a Mendelian randomization (MR) study conducted by Deng *et al*. ^24^ also found that the expression abundance of *RAB27B, GMPPB*, and *TMEM106B* were associated with the depression risk at both protein and mRNA levels, suggesting a potential causal effect of these genes in depression. Considering the significant genetic correlation between depression and anxiety disorders (r_g_= 0.87) (**Supplementary Fig.S3**), we also compared the risk proteins identified in this study and proteins identified in our previous PWAS of anxiety disorders (our unpublished manuscript). Three proteins, including TMEM106B, RAB27B, and CTNND1, showed significant associations with both depression and anxiety disorders. Of note, these three proteins showed proteome-wide significant associations in both ROSMAP and Banner brain pQTL datasets (**Fig.5b, c**), consistent with the results of TWAS (**Fig.5a**). These data strongly suggested that *TMEM106B, CTNND1, GMPPB*, and *RAB27B* are promising candidates for depression.

### Colocalization analysis

To investigate if the GWAS and QTL signals were driven by the same causal variants, we conducted colocalization analysis^25^. Colocalization analysis using eQTL (PsychENCODE) and GWAS signals showed shared causal variants for 87 genes (PP4>0.70), including *AREL1, DFNA5, FURIN, P2RX7, TMEM106B*, etc (**Fig.7** and **Supplementary Table S9**). When we restricted colocalization analysis on transcriptome-wide significant genes (Bonferroni corrected *P* threshold = 2.86 × 10^−5^), 63 genes showed colocalization.

Colocalization of pQTL (ROSMAP) and GWAS signals identified 25 candidate proteins, including P2RX7 (PP4=0.994), TMEM106B (PP4=0.992), CNNM2 (PP4=0.987), EPHB2 (PP4=0.986), etc (**Supplementary Table S9**). Colocalization of pQTL (Banner dataset) and GWAS signals identified 17 proteins (**Supplementary Table S9**). Of note, 15 genes were supported in the colocalization analysis of all three QTL datasets, including *P2RX7, TMEM106B, CNNM2, CSE1L*, etc (**Supplementary Table S9**), strongly suggesting that these genes are authentic candidates for depression.

### Mendelian randomization analysis identified potential therapeutic targets

To identify new drug targets and to seek potential drug repurposing opportunities, we performed Mendelian randomization (MR) analysis by using genetic variants associated with gene expression or protein abundance of 1,263 actionable drug proteins (approved or in clinical stage drugs therapeutic targets) curated by Gaziano *et al*.^26^.

MR analysis using eQTL from BrainMeta v2 dataset and GWAS signals identified 9 promising actionable drug targets (Bonferroni corrected *P* threshold < 7.50 × 10^−5^), including *ESR2, P4HTM, CD40, EPHB2, PSMC3, GRIK2, PRKCD, FOLH1*, and *CRHR1* (**Fig.6** and **Supplementary Table S15**). MR analysis using pQTL (ROSMAP dataset) and GWAS associations identified 13 promising candidate proteins, including DAGLA, P4HTM, DAGLB, EPHB2, TAOK3, etc (**Supplementary Table S16**). MR analysis using pQTL (Banner dataset) and GWAS associations identified 8 proteins (**Supplementary Table S17**). Of note, DAGLA, P2RX7, and PSMB4 were supported in the MR analysis of both pQTL datasets. And P4HTM, CD40, EPHB2, and GRIK2 showed MR significance at both mRNA and protein levels, strongly suggesting that these genes are promising drug targets for depression.

**Fig. 6.**
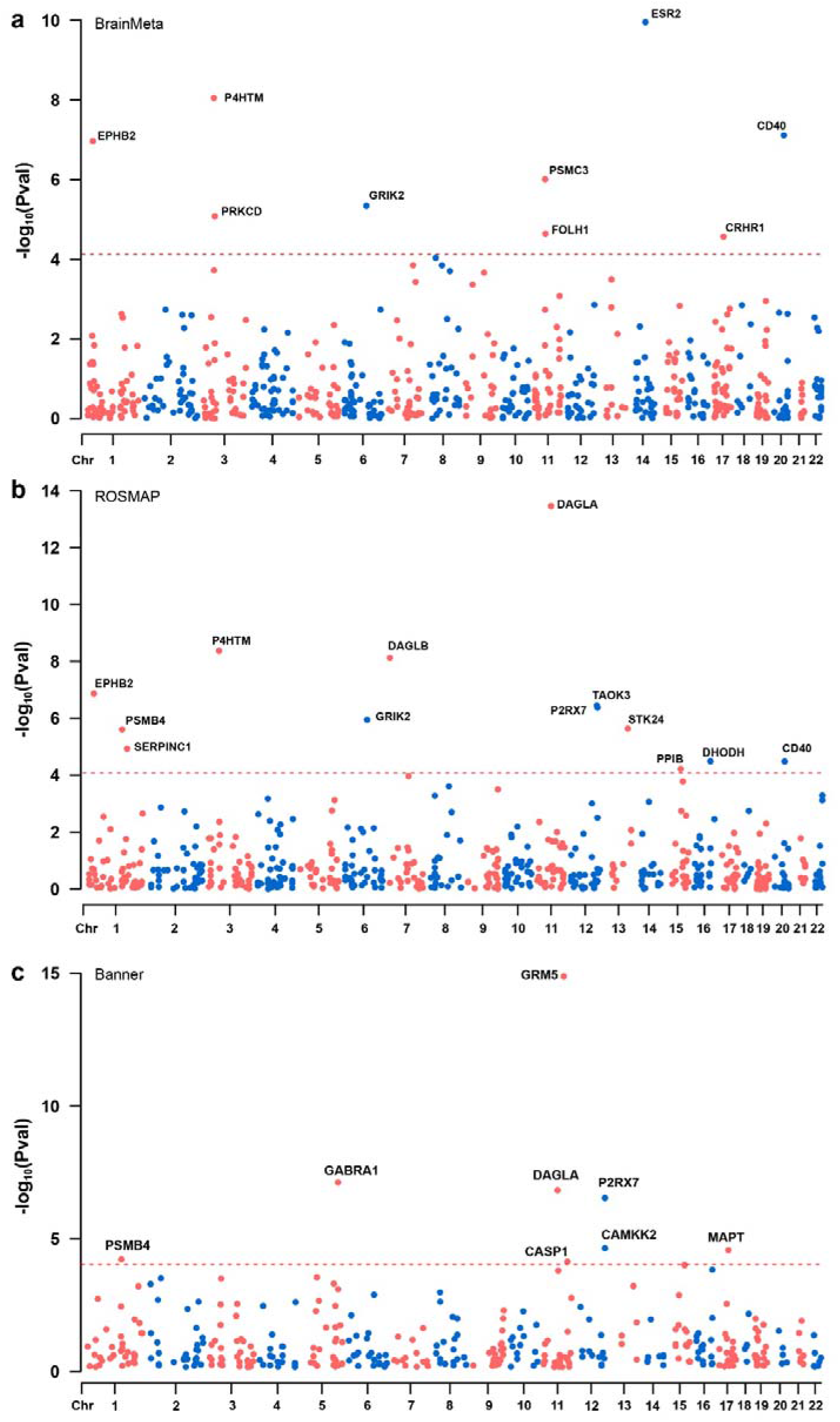
Drug target gene mendelian randomization analysis. Only MR significant genes (Bonferroni corrected *P* < 0.05) are shown. **(a)** BrainMeta eQTL MR results. **(b**,**c)** ROSMAP and Banner pQTL MR results.

### Heritability enrichment and gene set analysis

Tissue-based heritability enrichment analyses showed that the heritability of depression is mainly enriched in the brain tissues. Of note, depression associations showed the most significant enrichments in the cerebellar hemisphere and frontal cortex (BA9) (**Supplementary Fig.S5**). Significant enrichments were also observed in other brain regions (FDR < 0.05). Cell-type specific enrichment analysis showed significant enrichments in excitatory and inhibitory neurons, neuroblasts, and oligodendrocyte precursor cells (Bonferroni corrected *P* threshold < 1.28 × 10^−3^). These results identified neurons, neuroblasts, and oligodendrocyte precursor cells as possible responsible cell types for depression (**Supplementary Fig.S6**).

We subsequently used MAGMA to carry out gene set enrichment analysis and identified 12 significant enriched Gene Ontology (GO) terms. Of note, synaptic and postsynaptic membranes showed the most significant enrichment. In addition, the branching morphogenesis of nerves also showed significant enrichment (**Supplementary Fig.S7**). Taken together, these results identified neurons and synapses as the major cells and components for depression.

### Integrating ten lines of evidence prioritized most likely causal genes

To identify the most likely causal genes, we conducted gene prioritization by combing the evidence from 10 different analyses, including genomic location, TWAS, PWAS, colocalization, PoPS, functional genomic analysis, eQTL analysis, and SMR. The more lines of evidence support a gene, the higher probability that this gene is causal. A total of 34 genes were supported by at least four lines of evidence, of which 10 genes were supported by at least 7 lines of evidence (**Fig.7**). These genes include *TMEM106B, CTNND1, EPHB2, AREL1, CSE1L, RAB27B, STAU1, TMEM258, DCC, and PCDHA2. TMEM106B* was supported by all analyses and ranked the highest among these genes, strongly suggesting the causality of this gene. In addition, *CTNND1* was supported by 9 lines of evidence. *EPHB2* and *AREL1* were supported by 8 lines of evidence. Of note, *EPHB2* was also prioritized as a potential therapeutic target for depression in drug MR analysis. These results prioritized the most likely causal genes for depression.

**Fig. 7.**
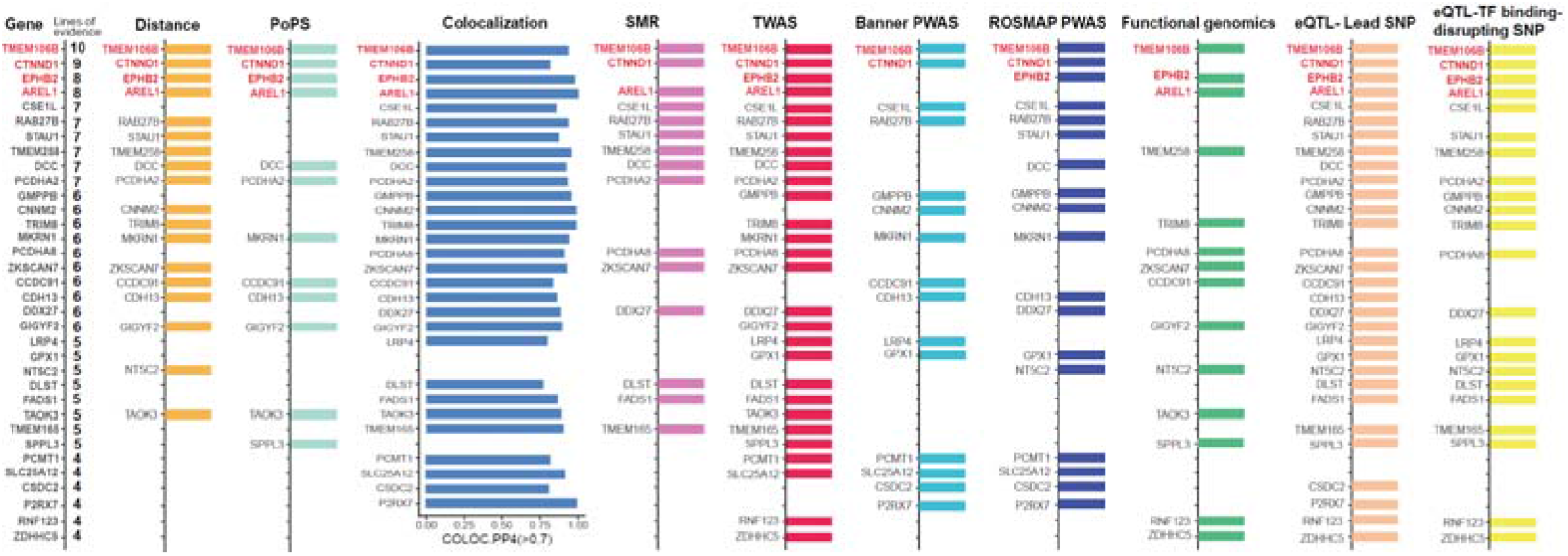
Gene prioritization results. Genes supported by at least 4 lines of evidence are shown (leftmost panel). Each color represents a type of supportive evidence, and genes supported by each type of evidence are shown. The 8 lines of causal evidence are as follows: (1) **Distance evidence**. For each risk locus, the gene located nearest to the reported lead SNP is considered to be the most possible candidate risk gene. (2) **PoPS evidence**. For each risk locus, PoPS was used to prioritize all potential causal genes, and the gene with the highest PoPS score was prioritized as the most possible candidate risk gene of this loci. (3) **Colocalization evidence**. For a specific gene, if eQTL signals of this gene and GWAS signals co-localized, this gene is listed as a potential risk gene (i.e., this gene was supported by colocalization analysis). (4) **SMR evidence**. Genes that showed SMR significance were considered as potential causal genes. (5) **TWAS evidence**. Genes showed transcriptome-wide significant associations were considered as potential causal genes. (6) **PWAS evidence (Banner dataset)**. Proteins whose expression levels were associated with depression were considered as potential causal genes. (7) **PWAS evidence (ROSMAP dataset)**. Proteins whose expression levels were associated with depression were considered as potential causal genes. (8) **Functional genomics evidence**. Functional genomics identifies risk SNPs affected by the binding of transcription factors. Genes located nearest the identified TF binding-disrupting SNPs are considered to be causal. (9) **eQTL evidence from lead SNPs**. Genes whose expressions are associated with the lead SNPs are considered as potential causal genes. (10) **eQTL evidence from TF binding-affecting evidence**. Genes whose expressions are associated with the TF binding-affecting SNPs are considered to be causal. *TMEM106B* was supported by all of the analyses. *CTNND1, EPHB2*, and *AREL1* were supported by more than seven lines of evidence.

### Enrichment of depression risk gene in synaptic process

Our gene prioritization analysis identified 34 high-confidence depression risk genes (Supported by at least four lines of evidence, see **Fig.7**). We used SynGO annotations^27^ to assess the role of these genes in synapses. Genes encoding postsynaptic specialization (*CTNND1, RAB27B*, and *EPHB2* gene) and presynapse ontology terms (*ZDHHC5, CTNND1*, and *DCC*) showed significant enrichment (**Supplementary Fig.S10** and **Supplementary Table S22**). Eight genes were mapped to SynGO biological processes annotations, involving synaptic signaling, synapse organization, and postsynapse and presynapse process (**Supplementary Fig.S11** and **Supplementary Table S22**). These results indicate significant enrichment of depression risk genes in synaptic process.

### Tmem106b knockdown resulted in depression-like behaviors

Animal model is pivotal to validate if risk genes identified by human genetic studies are involved in disease pathogenesis. Our above analyses indicated that *TMEM106B* ranked as the most possible risk gene for depression (**Fig.7**). To further investigate if *TMEM106B* is a *bona fide* depression risk gene, we knocked down *Tmem106b* in mouse hippocampus (a region with critical roles in depression)^28-30^ and performed serial behavioral experiment s(**Fig.8a-d**). Open field test showed that *Tmem106b* knockdown resulted in anxiety-like behaviors **(Fig.8e)**. light-dark transition assays showed that *Tmem106b*-knockdown mice moved less distance in the light box and disliked to shuttle between light and dark box compared with controls **(Fig.8f)**. Elevated plus maze test showed that the *Tmem106b*-knockdown mice preferred to stay in close arm (but not open arm) compared with controls **(Fig.8g)**. These results indicated that *Tmem106b* knockdown resulted in anxiety-like behaviors. We also evaluates the spatial working memory using the Y maze test, and found that the spatial working memory of *Tmem106b*-knockdown mice did not show a significant difference from controls **(Fig.8h)**. We next assessed depressive-like behaviors using the sucrose preference and tail suspension tests. We found that the sucrose preference of *Tmem106b*-knockdown mice was significantly decreased compared with controls **(Fig.8h)**. Besides, the immobility time of *Tmem106b*-knockdown mice was also significantly increased compared with controls **(Fig.8j)**. These results indicated that *Tmem106b* knockdown resulted in depression-like behaviors. Taken together, these behavioral results indicated that knockdown of *Tmem106b* in the ventral hippocampus can lead to anxiety- and depression-like behaviors in mice. The recapitulation of depression-like behaviors in *Tmem106b*-knockdwon mice provides robust evidence that supports our genetic findings, i.e., *TMEM106B* is a risk gene for depression.

**Fig. 8.**
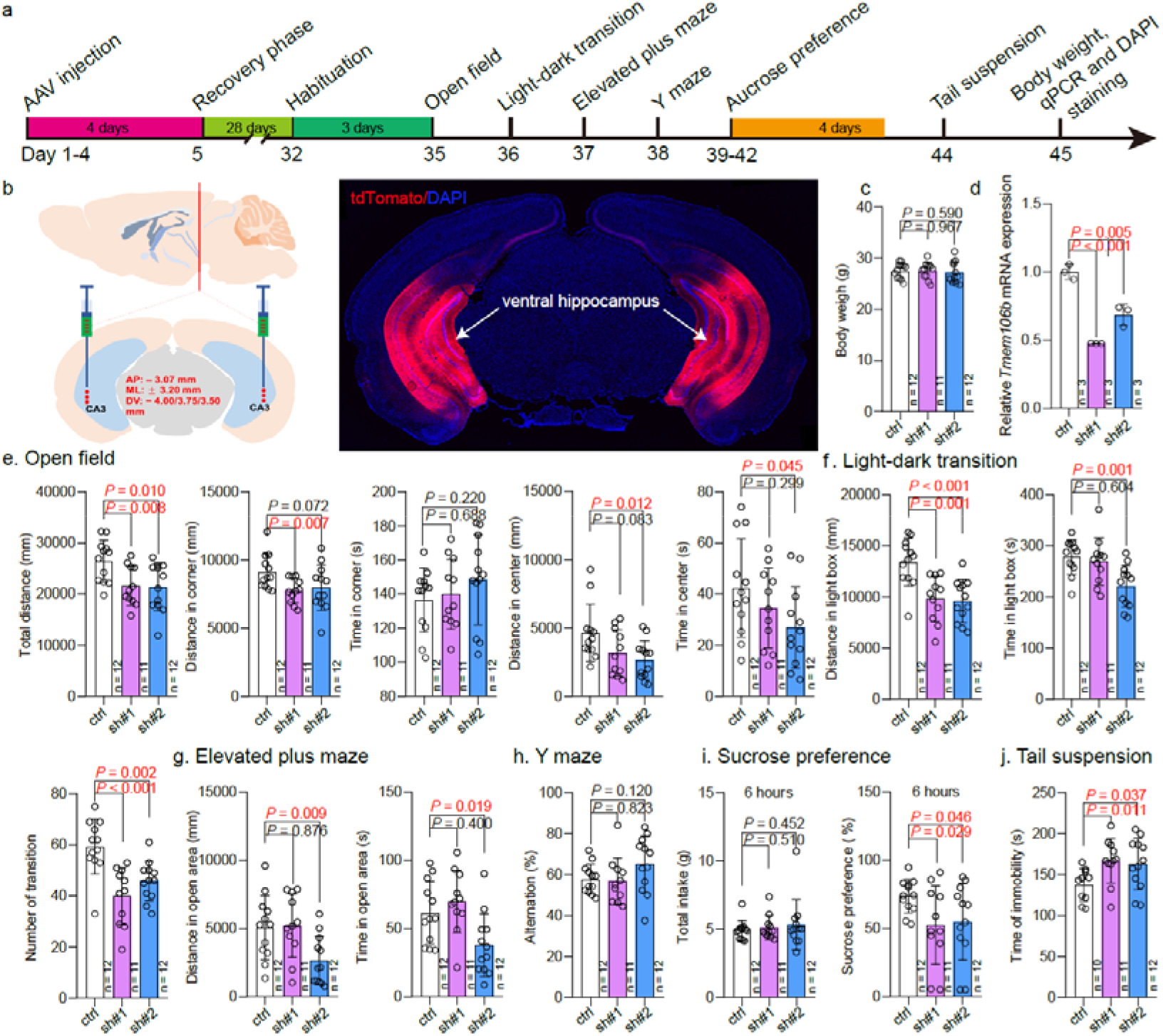
Tmem106b down-regulation in ventral hippocampus causes anxious and depressive behaviors in mice. (**a**) Schematic diagram of stereotactic injection and behavioral tests. (**b**) Left, schematic illustration of stereotaxic injection location in mouse ventral hippocampus; right, representative immunofluorescence image showed expression of tdTomato protein. shRNAs targeting *Tmem106b* were inserted into pAAV-CAG-tdTomato vector at NdeI site. Thus, expression of tdTomato indicated that the recombinant vectors were successfully injected to target sites. (**c**) The body weight did not show significant difference in *Temem106b-knockdown* and control mice. (**d**) qPCR showed that *Tmem106b* expression in ventral hippocampus was significantly down-regulated in *Tmem106b-knockdwon* mice compared with controls. Two pairs of complementary small hairpin RNA (shRNA) targeting *Tmem106b* were designed, and the first pair of shRNA (sh#1) showed higher knockdown efficiency. (**e**) Results of open field test. Total distance moved, distance moved in corner, time spent in corner, distance moved in center, and time spent in center area were analyzed. The results showed that total moved distance was significantly decreased in both *Tmem106b-knockdown* groups (shRNA#1 and #2) compared with control group. Distance moved in corner was significantly decreased in *Tmem106b-knockdown* (shRNA#1) mice compared with controls. Distance moved in center was also decreased *Tmem106b-knockdown* (shRNA#2) mice compared with controls. (**f-g**) Light-dark transition and elevated plus maze tests indicated that *Tmem106b* knockdown resulted in anxiety-like behaviors. (**f**) The *Tmem106b-knockdown* mice (shRNA#1 and #2) moved less distance in the light box compared with controls. Besides, the *Tmem106b-knockdown* mice (shRNA#2) disliked to shuttle between light and dark box compared with controls. (**g**) Elevated plus maze test showed that *Tmem106b-knockdown* mice preferred to stay in close arm than open arm compared with controls. (**h**) Y maze test showed that the spatial working memory of *Tmem106b-knockdown* and control mice did now show significant difference. (**i-j**) Depression-like behaviors were detected in sucrose preference test and tail suspension test. (**i**) Sucrose preference test showed that there was no significant difference in total intake (water and sucrose) in *Tmem106b-knockdown* and control mice. However, sucrose preference was significantly decreased in *Tmem106b-knockdown* groups compared with control group. (**j**) Tail suspension test showed that the immobility time was significantly increased in *Tmem106b-knockdwon* mice compared with controls. Two-tailed *Student’s t-test* was used for statistical test. Significance threshold was set at *P* < 0.05. Data represent mean ± SD. The sample size used in each test are as follows: for (**c, e-i**), n (ctrl) = 12, n (sh#1) = 11 and n (sh#2) = 12; for (**d**), n (ctrl) = 3, n (sh#1) = 3 and n (sh#2) = 3; for (**j**), n (ctrl) = 10, n (sh#1) = 11 and n (sh#2) = 12.

## Discussion

In this study, we carried out, to the best of our knowledge, the largest depression GWAS meta-analysis to date. By including 416,437 depression cases and 1,308,758 healthy controls, we identified 287 risk loci, of which 140 are new. In addition to reporting multiple novel risk loci for depression, we also performed a systems-level in-depth analysis to prioritize the causal variants and genes. For the first time, we generated the landscape of potentially causal genes for depression, which provides important opportunities to elucidate pathophysiology and develop new therapeutic targets.

One of the strengths of this study is the use of multi-ancestry cohorts. Considering most genetic studies of depression have been reported in European populations, it is of great importance to include other populations. In this study, with the use of European, Asian, and African ancestries, we identified 140 new risk loci for depression. Our cross-ancestry GWAS not only expands the risk loci of depression substantially but also provides important insights into genetic architecture of depression.

The lead SNP rs7531118 showed the most significant association with depression (*P* = 3.20 × 10^−31^) in our meta-analysis. Interestingly, this SNP is located near the *NEGR1*, a gene that has been frequently reported to be associated with depression^3,15,24,31,32^. *NEGR1* encodes neuronal growth regulator 1, a cell adhesion molecule of the immunoglobulin LON family. *NEGR1* has been reported to regulate neuronal migration and spine density during mouse cortical development^33^. In addition, Noh *et al*. found that *NEGR1* has a critical role in hippocampal neurogenesis, and loss of *NEGR1* resulted in anxiety- and depression-like behaviors in mice^32^. Interestingly, Carboni *et al*. showed that antidepressant treatment altered *Negr1* expression significantly^34^, further supporting the role of *NEGR1* in depression. These genetic and animal findings strongly implicate that *NEGR1* may have a pivotal role in the pathophysiology of depression. Besides, risk variants near genes such as *SORCS3, DRD2, H2BC7, LRFN5, DCC, IP6K1, RAB27B*, etc., also showed signficiant associations.

In addition to identifying novel risk loci, we also nominated functional (or potentially causal) variants from GWS loci using functional genomic analysis. We identified 64 TF binding-affecting SNPs from 287 risk loci, and our reporter gene assays showed that 80% (50 out of 62 tested SNPs) of the identified functional SNPs have regulatory effect, i.e., different alleles of these functional SNPs conferred significant luciferase activity differences. In fact, this is the most comprehensive and systematic functional genomic study to identify and elucidate the regulatory effect of depression risk variants. Considering the fact that identifying causal variants from risk loci is a major challenge in genetic study, the prioritization of functional variants provides an important starting point and opportunity to elucidate the regulatory mechanisms of depression risk variants.

Our findings indicate that these functional SNPs may exert their biological effect through alerting their binding affinity to TFs, and we dissected the regulatory mechanisms of the functional variants at single nucleotide level. Notably, the CTCF binding-affecting SNP rs7531118 (which is located near the *NEGR1*) also showed the most significant association with depression among the 64 TF binding-affecting risk SNPs, suggesting this functional SNP may confer depression risk by modulating *NEGR1* expression. Furthermore, *TMEM106B*, which is located 794 bp upstream of the rs1990622 (affects binding of CTCF), was supported by all lines of evidence in our gene prioritization analyses, strongly suggesting that the functional variant rs1990622 may confer depression risk by modulating *TMEM106B* expression.

Gene set enrichment analysis revealed that depression associations were significantly enriched in synapse-related cellular components and biological processes (**Supplementary Fig.S7**), suggesting the pivotal role of depression risk genes in synapse-related functions. Notably, some previous studies have also suggested that the synapse may have an important role in depression^8,35-39^. Our results further support that synapse dysfunction may play a key role in the etiology of depression.

Our drug-gene mendelian randomization analysis implicates potential therapeutic targets for depression, including *DAGLA, P2RX7, PSMB4, P4HTM, CD40, EPHB2*, and *GRIK2* (**Fig.6, Supplementary Table S15, S16, and S17**). Among these genes, *EPHB2, DAGLA, P2RX7*, and *PSMB4* are promising candidate targets. Among these genes, *EPHB2* was supported by 8 lines of evidence in our gene prioritization analysis (**Fig.7**), *DAGLA* reached the genome-wide significance level (**Supplementary Table S3**), *P2RX7* and *PSMB4* were supported by both PWAS and colocalization analysis (**Fig.5, Fig.7**). These findings indicated that these genes may represent promising therapeutic targets for depression. *EPHB2* encodes a member of the Eph receptor tyrosine kinase B subclass that has previously been shown to interact with NMDA receptors in excitatory synapses^40^. Evidence from cultured neurons and *EPHB2*-lacking mouse models showed that stimulation of Eph receptors modulates signaling pathways involved in synaptic plasticity and that lack of *EPHB2* leads to defects in synaptic plasticity in the mouse hippocampus^41^. In addition, studies have also found that the level of *EPHB2* and its downstream molecules are reduced in the medial prefrontal cortex (mPFC) of chronic social defeat stress mice^42^ (an animal model of depression), and *EPHB2* can participate in the regulation of stress-induced spine remodeling in mPFC^42,43^. *DAGLA* encodes diacylglycerol lipase alpha (DAGLa), which is involved in the biosynthesis of the endocannabinoid, 2-arachidonoyl-glycerol (2-AG)^44-46^. *DAGLA* knockout led to depression-related behaviors^47^. Furthermore, clinical studies have also shown that antagonists of DAGLa can increase the incidence and severity of depression^48,49^. In the brains of *DAGLA* knockout animals, 2-AG levels were significantly reduced (by 80%). Behavioral changes induced by loss of *DAGLA* include a range of classic depressive phenotypes, including decreased exploration of central areas of open space, increased behavioral hopelessness, and increased anxiety-related behaviors in light/dark boxes, etc. *P2RX7* encodes ATP-channel, purinergic receptor P2X, ligand-gated ion channel 7, P2RX7^50^. P2XR7 is primarily located in immune cells and glial cells of the central nervous system^51^. The role of dysregulation of the neuroimmune system, particularly the inflammatory response, in the development of symptoms of depression has been well described^52,53^. Of note, a previous study also nominated P2RX7 as a potential therapeutic target for depression, suggesting that P2RX7 receptor antagonists may represent potential drugs for depression^54-56^. *PSMB4* encodes proteasome 20S Subunit Beta 4. The 20S proteasome plays an important role in major histocompatibility complex (MHC) class I protein antigen peptide presentation^57^, which has been reported to be associated with susceptibility of depression in the previous genetic association studies^58^. Interestingly, *PSMB4* and *P2RX7* were also nominated as actionable drug targets for depression in our previous Mendelian randomization study^59^. These lines of evidence suggest that EPHB2, DAGLA, P2RX7, and PSMB4 are promising drug targets for depression. Further animal and clinical studies on these candidates will help to develop new therapeutic drugs for depression.

In addition to identifying risk loci and performing comprehensive integrative analysis, we also provided animal model evidence that supports *TMEM106B* is a risk gene for depression. Recaptitution of disease phenotypes is one of the most important and useful ways to validate if risk genes identified by genetic studies are involved in disease. We found that *Tmem106b* knockdown resulted in depression-like behaviors, indicating this gene has a critical role in depression. These animal model data provide robust evidence that support our GWAS findings.

In summary, we identified 140 novel risk loci and prioritized likely causal variants and genes for depression. More importantly, we validated the regulatory effect of most functional variants with reporter gene assays. Our study provides important insights into the genetic architecture of depression. Further functional characterization and mechanistic study of the identified risk genes will help to elucidate the neurobiology of depression and provide new therapeutics.

## Data Availability

The genome-wide summary statistics will be available from the corresponding author upon reasonable request.

## Acknowledgements

This study was equally supported by the Key Project of Yunnan Fundamental Research Projects (202101AS070055) and the Distinguished Young Scientists grant of the Yunnan Province (202001AV070006). We thank Miss. Qian Li for her technical assistance.

## Author contributions

XJL conceived, designed, and supervised the whole study. YFL, JYW, and SWL conducted the behavioral tests. RC performed reporter gene assays. XLD performed all the analyses, including meta-analysis, TWAS, PWAS, colocalization, fine mapping, SMR, and so on. XJL, YGY, ML, TL, and ZJZ contributed to this work in study design, data interpretation, and manuscript writing. XLD drafted the manuscript. XJL oversaw the project and finalized the manuscript. All authors revised the manuscript critically and approved the final version.

Brittany L. Mitchell

## Methods

### Genome-wide summary statistics used for meta-analysis

We performed an inverse variance-based fixed-effect meta-analysis by combining genome-wide associations from six large-scale, multi-ancestry GWAS, including MVP (83,810 cases and 166,405 controls of European ancestry, 25,843 cases and 33,757 controls of African ancestry)^3^, study from Howard et al. (23andme-UKB-PGC, 246,363 cases, and 561,190 controls)^5^, AGDS (13,318 cases and 12,684 controls)^9^, BioBank Japan (836 cases and 177,794 controls)^11^, East Asian ancestry (12,455 cases and 85,548 controls)^10^, and FINNGEN (33,812 cases and 271,380 controls) (https://www.finngen.fi/en)^60^. Briefly, for MVP, cases were defined using an ICD code-based algorithm to determine depression case status. Cases with at least one MDD inpatient diagnosis code or two MDD outpatient diagnosis codes were included in GWAS^3,61^. For 23andme-UKB-PGC dataset, a broad definition of depression was used for UK Biobank and more detailed phenotypic information can be found in the original paper^62^. For 23andMe data, samples were classified based on responses to web surveys, and those individuals who self-reported receiving a clinical diagnosis or treatment of depression were included as cases. For PGC samples, Wray *et al*. conducted a PGC cohort of European ancestry, focusing on obtaining the clinically derived phenotype of depression, which has been described previously^4^. The sample of AGDS dataset is from Australia Genetics of Compression Study, participants met the DSM-5 MDD standard (lifelong MDD) at some time in their lives but were not diagnosed as SCZ, BIP, or ADHD. More detailed information can be found in the original publication^9,63^. For BioBank Japan dataset, Sakaue *et al*. combined the past medical history and text mining of electronic medical records to conduct 220 deep-phenotype genome-wide association studies (including diseases, biomarkers, and drug use) in the Japanese Biological Bank. The dataset of depression was downloaded from https://pheweb.jp/download/Depression. For East Asian ancestry data, the samples from China Kadoorie Biobank (CKB), CONVERGE, Taiwan MDD research and the East Asian DNA samples of 23andMe companies in the United States and the United Kingdom were included. The details of each cohort can be found in the original article^10^. For FINNGEN, cases were assessed using ICD criteria. Cases that met unipolar depression (FINNGEN code: F5_DEPRESSIO, ICD-10 code: F32, F33) were included.

### Data processing and meta-analysis of genome-wide association study summary statistics

As several analytic methods, including logistic regression, linear regression, and logistic mixed models, were used in the original studies, we processed each dataset separately to obtain a unified data format for meta-analysis. For results obtained using logistic regression, effect size (beta) was converted into odds ratio using OR=exp^(Beta)^. We further processed each dataset to ensure the OR was based on the same effect allele. Besides, the standard error of ln(OR) and the corresponding *P* value were also calculated for all datasets. Finally, only variants with a minor allele frequency (MAF) > 0.01 were included in the meta-analysis.

To maximize discovery power^64^, we performed meta-analysis using an inverse variance-based fixed-effects meta-analysis implemented in PLINK (v1.09)^65^, which conducts meta-analysis based on the standard error weighted method. The combined effect size of the meta-analysis is the weighted average of individual study effect sizes (i.e., OR), which was calculated based on the logarithm of the effect size (OR) of each study and the inverse of the within-study standard errors (SE) of the effect size.

FUMA v1.3.7 was used to define the risk loci^66^, with the default parameters. Briefly, FUMA first defines independent lead SNPs (*P* < 5 × 10^−8^, r^2^ < 0.1), then defines risk loci by merging physically close or overlapping SNPs, lead SNPs that are closer than 250 kb were merged into one genomic risk locus. LD information was calculated using the 1000G phase3 EUR reference panel.

### LD score regression

Linkage disequilibrium Score regression (LDSC, https://github.com/bulik/ldsc)^12^ was used to estimate the SNP-based heritability and pairwise genetic correlations between the six depression GWASs (MVP, 23me-UKB-PGC, AGDS, BioBank Japan, East Asian ancestry, and FINNGEN). To control the genomic inflation, we also calculate the LDSC regression intercepts and attenuation ratio for the six contributing cohorts and the final meta-analysis results **(Supplementary Table S3)**. LDSC quantifies the genetic variability (LD score) for each SNP marker by the degree of linkage disequilibrium (r^2^) between SNPs. Then, linear regression was used to fit the relationship between LD score and Chi-square statistic to determine whether there were confounding factors in the GWAS results.

Meanwhile, LDSC can analyze the genetic correlations between traits by replacing the Chi-square statistic with the product of the z-scores from two traits^67^. In this study, we reformat the GWAS summary statistics using the standard procedures (describe detailed in https://github.com/bulik/ldsc/wiki/Heritability-and-Genetic-Correlation) and use the --r_g_ command to calculate the genetic correlations of the pairwise depression GWAS datasets. Traits (or phenotypes) used for LDSC analysis are provided in **Supplementary Table S5**.

### Identification of potential causal variants using functional genomics

We used the functional genomics approach^14,15^ to identify the functional (or potential causal) variants from the identified risk loci. Briefly, chromatin immunoprecipitation sequencing (ChIP-Seq) assays that used brain tissues or neuroblastoma cell lines (a total of 30 transcription factors (TFs)) were downloaded and processed as previously described^14,15^. Based on these ChIP-Seq data, we derived the DNA binding motifs of these TFs. We further used FIMO^68^ to compare the derived binding motifs with the publicly available position weights matrix (PWM)^16^, and the best-matched motifs were used for further analysis. We extracted the SNPs that were in linkage-disequilibrium (LD) (r^2^ > 0.8) with the lead SNPs using the 1000 genome phase3 data of European population from the SNiPA website (https://snipa.helmholtz-muenchen.de/snipa3/)^69^. Finally, we investigated if the lead SNPs or SNPs in LD with the lead SNPs affect the binding of TFs. Detailed information about functional genomics can be found in previous studies^14-16^.

### Identification of potential causal variants using fine-mapping

To identify the potential causal variants, we carried out statistical fine-mapping using FINEMAP^17^ and SuSiE^18,19^. FINEMAP uses a shotgun stochastic search algorithm to prioritize a set of the most possible causal variants of the region. SuSiE prioritizes the possible causal variants based on a new model for sparse multiple regression which focuses on quantifying uncertainty in which variables should be selected. Both FINEMAP and SuSiE allow for multiple causal variants in a risk locus. To perform the fine-mapping analyses, we first extracted the SNPs in linkage disequilibrium (LD) (r^2^ ≥ 0.8) with the lead SNPs using European genotype data from the 1000 Genomes projects (Phase 3)^70^. For SuSiE fine-mapping, after extracting variants present in the linkage disequilibrium reference panel, we used the susie_rss() function with default settings in susieR while allowing up to 10 putative causal variants per locus (L=10). For FINEMAP fine-mapping, We first compute the LD matrix using the CalcLD_1KG_VCF.py package with PAINTOR^71^, and then performed fine-mapping analysis using --sss flag under the assumption of 3 causal variants per loci (--n-causal-snps 3). In total, we attempted to fine-map 287 GWS risk loci and retained results with a posterior probability (PIP) greater than 0.7 **(Supplementary Tables S10** and **S11)**.

### Expression quantitative trait loci analysis

To identify genes whose expression were associated with the risk variants, we examined the associations between the identified risk variants (including lead SNPs, functional SNPs identified by functional genomics, and causal variants prioritized by fine-mapping approaches) and gene expression in the human brain. Brain eQTL data from the BrainMeta v2 dataset^72^ (N=2,865) were used in this study. The BrainMeta v2 dataset contains eQTL results for brain cortex tissue from seven cohorts, including BrainGVEX, the Lieber Institute for Brain Development, the CommonMind Consortium, and the CommonMind Consortium’s National Institute of Mental Health Human Brain Collection Core, Mount Sinai Brain Bank (including four cortex regions: BM10, BM22, BM36, and BM44), Mayo Clinic and ROSMAP, detailed information can be found in the original publications^72-75^.

### Transcriptome-wide association study

To identify genes whose genetically-regulated expression changes are associated with depression, we performed a TWAS through FUSION package^22^ by integrating the depression GWAS summary statistics and brain eQTL data (N=1,321) from PsychENCODE^21^ with default settings. TWAS analysis utilizes several linear models (including BLUP, BSLMM, LASSO, Elastic Net, and top SNPs) to calculate SNP expression weights representing the association between SNPs and gene expression in a reference set. The SNP-expression weights of PsychENCODE used in this study were downloaded from http://resource.psychencode.org/. Detailed information on the calculation of SNP-expression weights is available on the PsychENCODE website and related publications^21^. Finally, the Z-score result calculated by TWAS was used to evaluate the association between genes and depression, and the absolute value of Z-score reflected the strength of the association between risk genes and diseases. Significance thresholds for TWAS results were adjusted using the Bonferroni correction (corrected *P* threshold = 2.86 × 10^−5^). Genes that passed the Bonferroni correction were considered statistically significant TWAS results.

### Proteome-Wide Association Study

To identify proteins whose genetically-regulated abundance is associated with depression, we conducted PWAS by integrating genome-wide meta-analysis results and two independent protein quantitative trait loci (pQTL) datasets from the dorsolateral prefrontal cortex (ROSMAP: n=376, Banner: n=152)^23^. Briefly, Wingo *et al*. conducted a proteomic study and used FUSION to estimate protein weights in discovery and confirmation datasets separately. We downloaded the processed protein weight files from https://www.synapse.org/ (Synapse ID: syn9884314 and syn23245237) and performed PWAS analysis as described in TWAS.

### Colocalization analysis

To explore whether the eQTL (from PsychEnCODE^21^), pQTL (from the Banner and ROSMAP^23^) and GWAS signals were co-localized, we performed colocalization analysis with Coloc package^25^ implemented in FUSION^22^. Only genes with corrected TWAS or PWAS *P* < 0.05 were included for colocalization analysis.

### Drug-gene mendelian randomization analysis

For the Drug-gene MR analysis, the TwoSampleMR R package was used to perform two-sample MR analysis (v0.5.6, https://mrcieu.github.io/TwoSampleMR/)^76^. We used 1263 actionable druggable genes selected by Gaziano *et al*. as potential candidates in this study^26^. To identify drug re-purposing opportunities, Gaziano *et al*. used data from ChEMBL (version 26) to identify 1,263 human proteins as actionable (i.e., the approved or clinical-stage drugs therapeutic targets) drug targets ^26,77^. More detailed information on these actionable drug targets can be found in the study by Gaziano *et al*^26^. Associations between genetic variants and gene or protein expression abundance (i.e., eQTL and pQTL) were used as exposure instruments, and the meta-analysis result was used as outcome data. The brain eQTL data was from the BrainMeta v2 dataset^72^, and the brain pQTL data was from the Banner and ROSMAP datasets^78^. Considering the sample size of the pQTL analysis was small, we used a relatively relaxed *P* threshold (i.e., 0.05) for pQTL datasets, as described in previous studies^79-81^, and used a more stringent *P* value threshold (i.e., 5×10^−8^) for the eQTL dataset. Subsequently, actionable druggable genes from pQTL and eQTL results were obtained as QTL data sets for the MR analysis. Independent SNPs were defined if the linkage disequilibrium value (r^2^) was less than 0.001. Then, the exposure and outcome data were harmonized using harmonise_data() function to ensure the same effect allele in exposure and outcome data. For exposures with only one IV, the Wald ratio method was used, and for exposures with two or more IVs, the inverse variance weighted (IVW) method was utilized.

### Summary-data-based Mendelian Randomization

We used the Summary-data-based Mendelian Randomization (SMR) method^82^ (which integrates GWAS and gene expression quantitative trait loci (eQTL) data) to identify causal genes associated with depression. The cis-eQTL data used as instrumental variables (IVs) for gene expression were from the PsychENCODE project (n = 1387)^83^ and were downloaded from the SMR website (https://yanglab.westlake.edu.cn/data/SMR/PsychENCODE_cis_eqtl_HCP100_summary.tar.gz). We ran SMR analysis with default parameters (--peqtl-smr 5.0e-8, --maf 0.01, --peqtl-heidi 1.57e-3, --heidi-mtd 1). Heterogeneity tests were conducted with the HEIDI test. We excluded results with significant heterogeneity (P_HEIDI_ > 0.01) from the SMR results and used the Bonferroni-corrected significance threshold (corrected *P* threshold = 4.88 × 10^−6^) to identify causal genes that reach SMR significance.

### Tissue and cell type enrichment analysis of depression heritability

To explore whether the genome-wide associations of depression were enriched in specific tissues or cell types, we performed heritability enrichment analysis using MAGMA (v1.1.0)^84^. For the tissue enrichment analysis, we used gene expression data (from the Genotype-Tissue Expression (GTEx) consortium, GTEx_Analysis_2017-06-05_v8_RNASeQCv1.1.9) of different human tissues to identify genes that are expressed in specific tissues. Based on the GWAS *P*-value, MAGMA uses a multiple linear principal component regression model to quantify the degree of association between genes and depression. MAGMA then tested whether these genes were enriched among genes specifically expressed in specific tissues. Cell type enrichment analysis was also performed using MAGMA. The single-cell RNA sequencing data that we used for cell type enrichment analysis was downloaded from Bryois et al. ^85^(https://github.com/jbryois/scRNA_disease/tree/master/Code_Paper/Code_Zeisel), A detailed description of this dataset can be found in the previous publications^86,87^. *P*-values for MAGMA enrichment analyses were corrected for the false discovery rate (FDR) using the p.adjust() function from R4.1.1.

### Gene set enrichment analysis

Gene set analysis was performed using MAGMA software. First, MAGMA maps SNPs to genes by using GWAS summary statistics and gene annotation files. Then, MAGMA calculated the association between specific genes and depression. Finally, MAGMA performs gene set enrichment analysis by testing whether the target gene set is more associated with depression than other genes that are not included in the gene set. *P*-values for competing gene sets were used to determine the level of significance. We downloaded all GO terms (including cellular components, biological processes, and molecular functions) and KEGG pathway gene sets from the MSigDB database (https://www.gsea-msigdb.org/gsea/msigdb/human/collections.jsp#C5). The final *P*-value of the MAGMA gene set enrichment analysis was corrected for the false discovery rate (FDR) using the p.adjust() function from R4.1.1.

### Polygenic Priority Score (PoPS) prioritization of candidate causal genes

We used the PoPS method to prioritize the genes near all SNPs in highly LD with the lead SNP (r^2^ > 0.8). PoPS is a gene prioritization method^88^, which uses the whole-genome signals in the GWAS summary statistical data, and combines a large number of bulk and single-cell expression data sets, biological pathways, and predicted protein-protein interaction data to prioritize candidate causal genes. We used the gene list and feature files downloaded from the PoPS GitHub page (https://github.com/FinucaneLab/pops) to calculate the PoPS score of all candidate genes. For each locus, only the gene with the highest PoPS score will be marked as the final candidate gene.

### Integrating different lines of evidence for gene prioritization

To prioritize the most possible causal genes for each risk locus, we combined evidence from 10 different analyses, including gene location, PoPS, colocalization, TWAS, PWAS, functional genomics, eQTL, and SMR. For simplicity, we assigned one point to each line of evidence, and the total score of each gene was the sum of all lines of evidence. Higher scores indicate a higher probability of being causal.

### Enrichment of depression risk gene in synaptic process

We tested the enrichment of 34 prioritized depression risk genes in the synaptic-related process (biological processes and cellular components) using SynGO (SynGO release: 20210225)^27^. The background gene set was used as the default “brain expression”, the evidence filters were set to “Medium stringency”, and the minimum number of genes for each term in the GSEA analysis was set to 3. For more detailed content, see the SynGO website description (https://syngoportal.org/help).

### Reporter gene assays

Reporter gene assays were performed as previously described^14,89,90^. The DNA sequences (approximately 600 bp) containing the candidate functional SNPs were inserted into the pGL4.11 vector, which is a basic vector without a promoter. Thus, this vector can be used to test the regulatory activity of SNPs located in promoter regions (referred as promoter assays). For SNPs that are not located in promoter region, DNA sequences containing the test SNPs were inserted into pGL3-Promoter vectors to detect enhancer activity (enhancer assays). All the inserted DNA sequences were verified by Sanger sequencing.

The human neuroblastoma cells (SH-SY5Y), which were originally obtained from the Kunming Cell Bank, Kunming Institute of Zoology, were used for reporter gene assays. Polymerase chain reaction (PCR) was conducted periodically to detect mycoplasma during cell culture to ensure no mycoplasma contamination. High-glucose Dulbecco’s Modified Eagle’s Medium (DMEM) supplemented with 10% fetal bovine serum (FBS) (Gibco, Cat. No: 10091148), 1% penicillin and streptomycin (100 U/mL), 10 mM sodium pyruvate solution (Gibco, Cat. No: 11360070), 1 × Minimum Essential Medium non-essential amino acid solution (Gibco, Cat. No: 11140050) was used for cell culture. All cells were cultured in a cell culture incubator with 5% CO_2_ at 37 °C.

For cell transfection and reporter gene detection, the constructed vectors (150 ng) and the internal control vector pRL-TK (Promega, Cat. No: E2241) (50 ng) were co-transfected into the SH-SY5Y cells using Lipofectamine™ 3000 (Invitrogen, Cat. No: L3000-015). SH-SY5Y cells were plated into 96-well plates containing 100 μl medium at densities of 6.0 × 10^4^ cells/well. After forty-eight hours post transfection, a dual-luciferase reporter gene assay system (Promega, Cat. No: E1960) was used to measure the luciferase activity. Differences were calculated with two-tailed *Student’s t test*, and the significance threshold was set at *P* < 0.05.

### Behavioral assays

#### Construction of Tmem106b knockdown plasmid

To knock down *Tmem106b* expression, we designed two shRNAs (shRNA#1:5’-GCGTTACATCGACAGACAA-3’; shRNA#2:5’-GGAATTTACTGGAAGAGAT-3’) targeting mouse *Tmem106b* exon-3. Non-specific shRNA sequence (5’-GATTTGCTGTTCGCCCAAG-3’) was used as negative control. The DNA sequences (containing U6 promoter and shRNAs) were synthesized and inserted into pAAV-CAG-tdTomato (Plasmid #59462, Addgene) at NdeI site, and the recombinant vectors (pAAV-U6-shRNA#c/1/2-CAG-tdTomato) were purchased from TSINGKE Biological Technology (TSINGKE, Co., Ltd, Beijing, China).

#### Adeno-Associated Virus (AAV) production and purification

HEK293T cells with 80-90% confluence (cultured in 15 cm cell-culture dishes) were used to produce AAVs. 10 ug recombinant vectors and AAV packaging plasmids (10 ug AAV-DJ and 20 ug pHelper) were co-transfected into HEK293T cells using polyethyleneimine (PEI) method. 72 hours post-transfection, cells containing target AAVs were collected and resuspended with cell lysis buffer (0.15M NaCl; 20mM Tris-HCl) and subjected to 4 times freeze-thaw cycles (between liquid nitrogen and 37 °C water) to release AAVs. 0.5% sodium deoxycholate and 50 U/mL benzonase (Cat. No: E1014, Sigma) were added into above cell lysis buffers to digest non-viral DNA at 37 °C for 30 min, then centrifugated by 7000 × rpm for 90 mins at 4 °C. The supernatants containing target AAVs were then subjected to gradient centrifugation using 15%, 25%, 40%, 54% iodixanol (Cat. No: D1556, Sigma-Aldrich) under 59,000 × rpm for 3 hours at 18 °C. The target AAVs were centrifugated into 40% iodixanol, and then collected the 40% iodixanol for further purification.

The iodixanol was removed by using Amicon ultrafiltration tube (Cat. No: UFC910096, Millipore) and 50 mL precool 1 × HBSS. Finally, 100-300 μL HBSS residues were used to suspend the purified target AAVs, and the collected AAVs were stored at -80°C. The virus titer was determined following the protocol of AAV Titration by qPCR Using SYBR Green Technology (https://www.addgene.org/protocols/aav-titration-qpcr-using-sybr-green-technology/)^91^.

#### Brain stereotactic injection

6-8 weeks C57BL/6J male mice were purchased from Institute of Model Animals, Nanjing University (Nanjing, China), and were raised in 12 h light/dark cycle SPF house with 23±2°C, 50-60% humidity. Mice were allowed to intake food and water freely. All animal experiments were approved by the Animal Ethic Committee of Kunming Institute of Zoology (IACUC-RE-2023-02-002).

Mice were adapted to new breeding environment for 1-2 weeks before brain stereotactic injection. Mice were anesthetized with isoflurane gas, then the head of mice was fixed on the brain stereotactic apparatus horizontally (RWD Life Science Co., Ltd.). Detailed information about viral injection is provided in **Supplementary material**.

#### Real time quantitative PCR (qPCR)

Total RNA was extracted from fresh hippocampal tissues with TRIzol Reagent (Cat. No: 10296028, Invitrogen). 1 μg total RNA was used as template to synthesize cDNAs using PrimeScript™ RT reagent Kit with gDNA Eraser (Cat. No: RR047A, Takara). The *Actb* and *Tmem106b* expression were quantified using TB Green™ Premix Ex Taq™ II (Tli RNaseH Plus) (Cat. No: RR820A, Takara) and CFX96 Touch™ Real-Time PCR Detection System (Bio-Rad). *Actb* was set as an internal gene to calculate relative gene expression using 2^^–ΔΔCt^ method^92^. Primers for *Actb* were 5’-GGCTGTATTCCCCTCCATCG-3’ (forward) and 5’-CCAGTTGGTAACAATGCCATGT-3’ (reverse), and primers for *Tmem106b* were 5’-AACATTGGCCCACTTGATATGAA-3’ (forward) and 5’-GAGTGTCCAAAGTATGCTGTTGT-3’ (reverse).

#### Behavioral tests

Six behavioral tests (open field, light-dark transition, elevated plus maze, Y maze, Sucrose preference and tail suspension) were performed to evaluate the depressive and anxious behaviors. All behavioral tests were conducted in an independent and undisturbed room between 10:00 a.m. and 6:00 p.m. About 24 hours interval was applied to minimize the stress effects. SuperMaze and SuperTst software (Shanghai XinRuan Information Technology Company) were used to record the test mice behaviors.

#### Open field test

Open field test is a method to evaluate movement and anxious behaviors of animals in novel environments. Open field test was conducted as previously described^93^. Briefly, the open field arena (Length: 40 cm; Width: 40 cm; Height: 40 cm) was divided into sixteen squares using SuperMaze software, the four squares at corner were defined as corner areas, and the four squares at center were defined as central areas. Each test mouse was placed in the same corner and allowed to freely explore novel environment for 5 min. Their movement tracks were recorded by video camera and SuperMaze software with the same parameters. The apparatus was cleaned with 10% ethanol before each trial. A total of 35 mice were tested successfully (control: n = 12; *Tmem106b*-shRNA#1: n = 11; *Tmem106b*-shRNA#2: n = 12). The time and distance of central and corner areas are indicators to reflect anxious levels.

#### Light-dark transition test

The light-dark transition test uses the conflict between innate aversion to light areas and exploratory behavior of rodents to study anxious behaviors of mice. Light-dark transition test was conducted as previously described^93^. Briefly, the apparatus consists of two same boxes (length: 20 cm; width: 15 cm; height: 25 cm), a light and dark box. The test mouse was placed in the center of the light box with head back to the hole, and allowed to freely shuttle between light and dark box for 10 mins. Their movement tracks in light box were recorded by video camera and SuperMaze software with the same parameters, and transition times were manually recorded. The apparatus was cleaned with 10% ethanol before each trial. A total of 35 mice were tested successfully (control: n = 12; *Tmem106b*-shRNA#1: n = 11; *Tmem106b*-shRNA#2: n = 12). Time and distance spent in light box and the transition times evaluated their anxiety levels.

#### Elevated plus maze test

Elevated plus maze investigates the anxiety state of animals based on the contradictory behaviors of exploring the new environment and the fear of hanging in open arms. Elevated plus maze was conducted as previously described^93^. Briefly, the elevated plus maze apparatus is above the ground 75 cm and consists of four arms (two close arms (length: 35 cm; width: 5 cm; height: 15 cm) and two open arms (length: 35 cm; width: 5 cm) and a cross area (length: 5; width: 5 cm). The test mouse was placed in cross area facing the same open arms. Their movement tracks in elevated plus maze apparatus were recorded by video camera and SuperMaze software with the same parameters. The apparatus was cleaned with 10% ethanol before each trial. A total of 35 mice were tested successfully (control: n = 12; *Tmem106b*-shRNA#1: n = 11; *Tmem106b*-shRNA#2: n = 12). Time and distance spent in the open arms were recorded to reflect anxiety levels.

#### Y maze test

Y-maze investigates spatial working memory in mice based on spontaneous tendency to explore new environments. Y maze test was conducted as previously described^93^. Briefly, Y-shaped geometric arena with three arms (arm length, width, height: 35 cm, 5 cm, 15 cm) were defined as A, B, and C arms, and the angle between arm is 120°. The test mouse was placed in A arm and permitted to freely explore Y maze apparatus for 8 mins. Their movement tracks in Y maze apparatus were recorded by video camera and SuperMaze software with the same parameters, and manually recorded the series of arm entries. The spontaneous alternation event was defined as three consecutive arm entries that are different. The apparatus was cleaned with 10% ethanol before each trial. A total of 35 mice were tested successfully (control: n = 12; *Tmem106b*-shRNA#1: n = 11; *Tmem106b*-shRNA#2: n = 12). Spontaneous alternation percent in Y-maze was evaluated their spatial working memory levels.

#### Sucrose preference test

Sucrose preference test utilizes rodents’ preference for sweetness to evaluate the anhedonia and depression of mice. Sucrose preference test was conducted as previously described^94^. Briefly, the test mouse was firstly fed in individual cage and adapted to two bottles (one containing water, the other containing 1% sucrose solution) for 48 hours, and the position of two bottles was changed at 24 hours to avoid position and bottle preference. Secondly, the two bottles were taken away for 24 hours after adaptation period. Finally, the mice were permitted to freely access water and sucrose solution for 6 hours and switched the positions of two bottles at 3 hours. The water and sucrose solution in each cage were measured before and after 6 hours. Sucrose preference (%) = sucrose consumption/(sucrose consumption + water consumption) × 100%. A total of 35 mice were tested successfully (control: n = 12; *Tmem106b*-shRNA#1: n = 11; *Tmem106b*-shRNA#2: n = 12). Sucrose preference was evaluated their anhedonia and depressive levels.

#### Tail suspension test

Tail suspension test was used to detect desire to survive of mouse in a desperate situation, the mouse frequently has no desire to survive in depressive state. Tail suspension test was conducted as previously described^93^. Briefly, mouse tail (about 1 cm) was fixed on the suspension hook in chamber box (length: 20 cm; width: 20 cm; heigh: 32 cm) using adhesive tape, and let the mouse body freely hung down in the air at a fixed height for 6 mins. Their activity (swing, struggle, and climb) were recorded by video camera and SuperTst software with the same parameters. A total of 33 mice were tested successfully (control: n = 10; *Tmem106b*-shRNA#1: n = 11; *Tmem106b*-shRNA#2: n = 12). Immobility state (swing) in the last 4 mins was used to evaluate their depressive levels.

