## Supplementary for "Cross-ancestry genome-wide association study and systems-level integrative analyses implicate new risk genes and therapeutic targets for depression"

**Supplementary methods**

***Brain stereotactic injection***

First, the hair of the mouse's head was removed using automatic razor for preparing longitudinal incision to expose the skull surface. Second, the anterior fontanelle of mouse was set as the origin (AP: 0 mm; ML: 0 mm; DV: 0 mm), and calibrated the horizontal plane of brain. Third, two symmetrical holes were drilled according to ventral hippocampal coordinates using an electric skull drill (RWD Life science Co., Ltd.). Forth, the glass injection needle inserting into micro-syringe (RWD Life science Co., Ltd.) was filled with target AAVs (titer=5×10^12^ GC/mL), and placed pointing to anterior fontanelle (which was set as origin). Fifth, the injection needle was placed to point to the target brain point (AP: – 3.07 mm; ML: ± 3.20 mm; DV: − 4.00 mm) and stayed five minutes. Sixth, the syringe pump controller (RWD Life science Co., Ltd.) parameters were set as follows: speed: 2 nL/s, volume: 500nL. Three coordinate points were injected (DV: − 4.00, − 3.75, − 3.5 mm). Finally, the scalp was sutured using absorbable surgical thread and sterilized the incision.

**Supplementary figures**

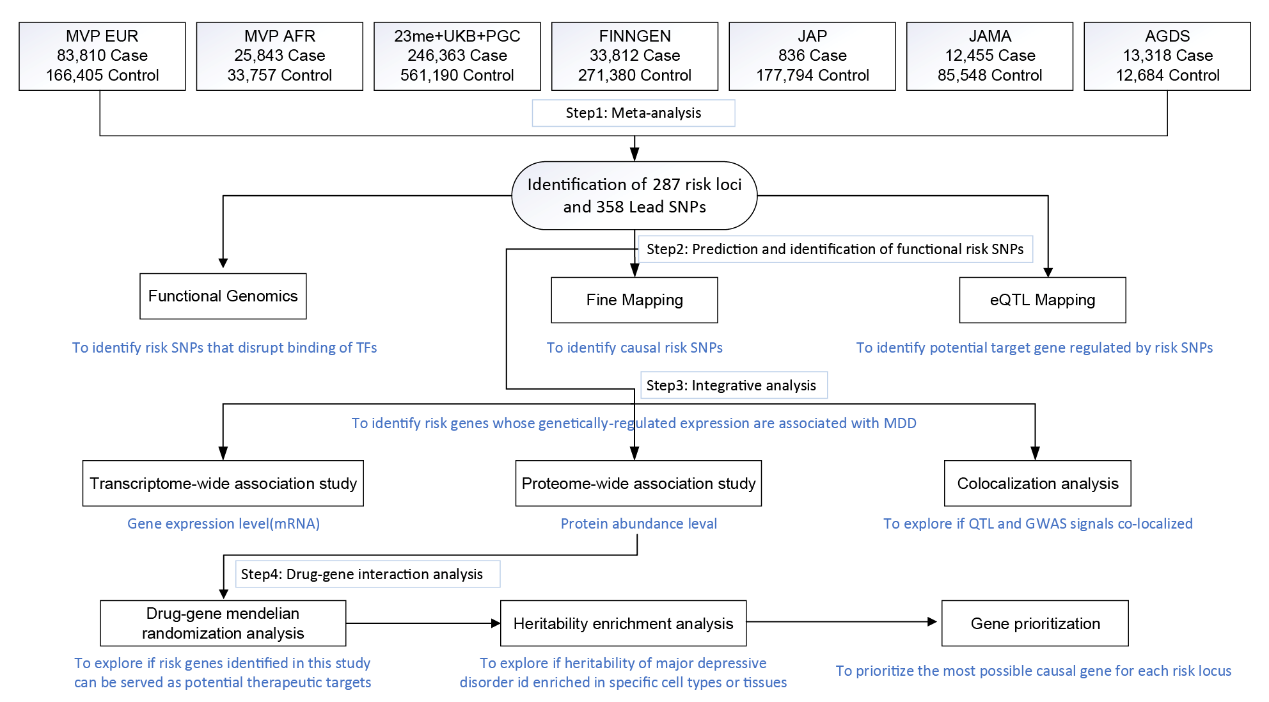

**Fig.S1. Overview of the analysis included in this study.** A meta-analysis was first conducted to identify novel risk loci for major depressive disorder. Based on the results of the meta-analysis, several analyses were then performed. SNP-based analyses, including fine-mapping, functional genomics, and expression quantitative trait loci (eQTL), were conducted to prioritize the functional (or potential causal) risk variants. Fine-mapping was conducted to prioritize the potential causal variants, functional genomics was performed to identify the SNPs that disrupt the binding of transcription factors (TFs), and eQTL was used to examine if the risk variants were associated with gene expression in the human brain. We further carried out gene-based analyses to prioritize the potential risk genes. Transcriptome-wide association study (TWAS) and proteome-wide association study (PWAS) were conducted to identify genes whose genetically-regulated expression is associated with major depressive disorder. Co-localization analysis was performed to explore if the QTL and GWAS signals were driven by the same variants. We conducted a drug-gene mendelian randomization analysis to identify potential therapeutic targets, and we investigated whether the heritability of major depressive disorder is enriched in specific tissues or cell types. Based on lines of evidence from the above analyses, we performed a gene prioritization analysis to prioritize the most possible risk genes for major depressive disorders. Finally, we explored the potential role of *Tmem106b* (the rat homolog of the human *TMEM106B* gene) in mouse behavior.

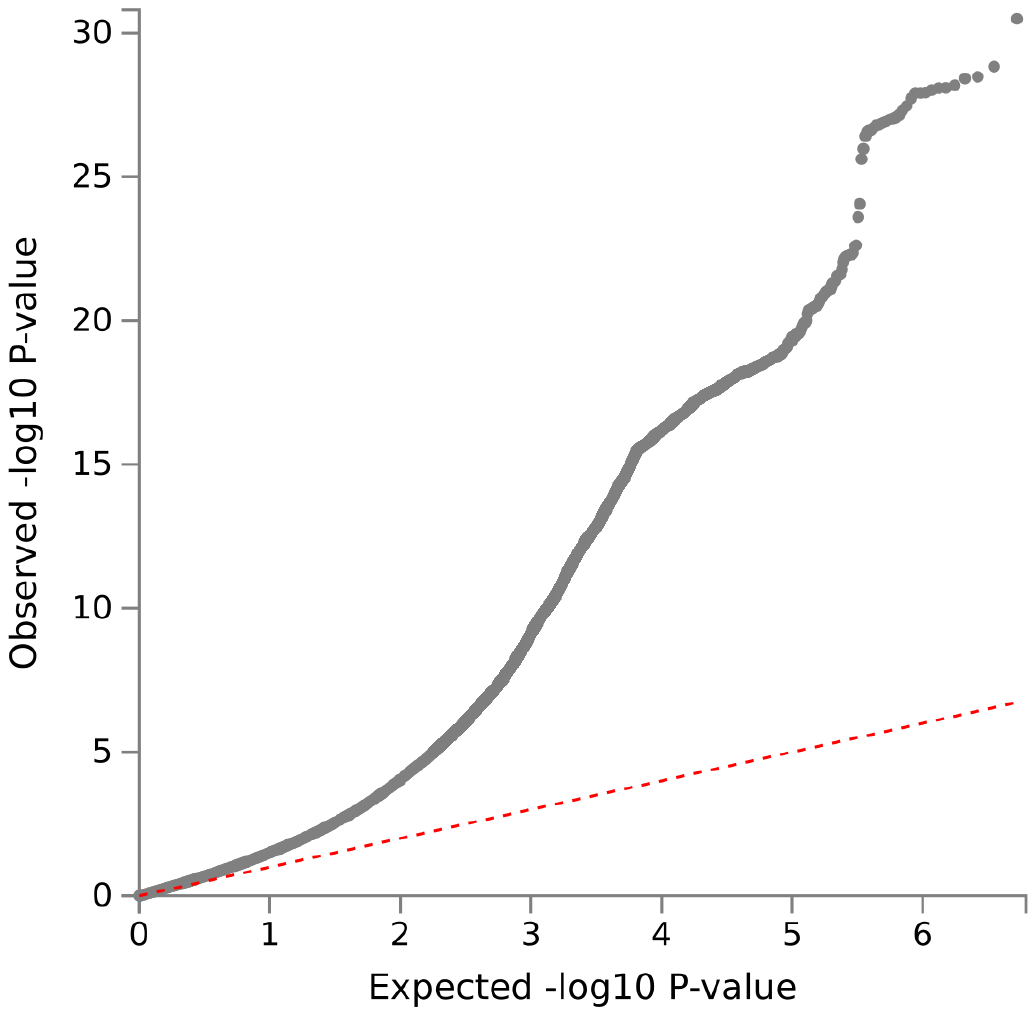

**Fig. S2.** **Quantile-quantile Plot Illustrating the GWAS Meta-Analysis for major depressive disorder.**

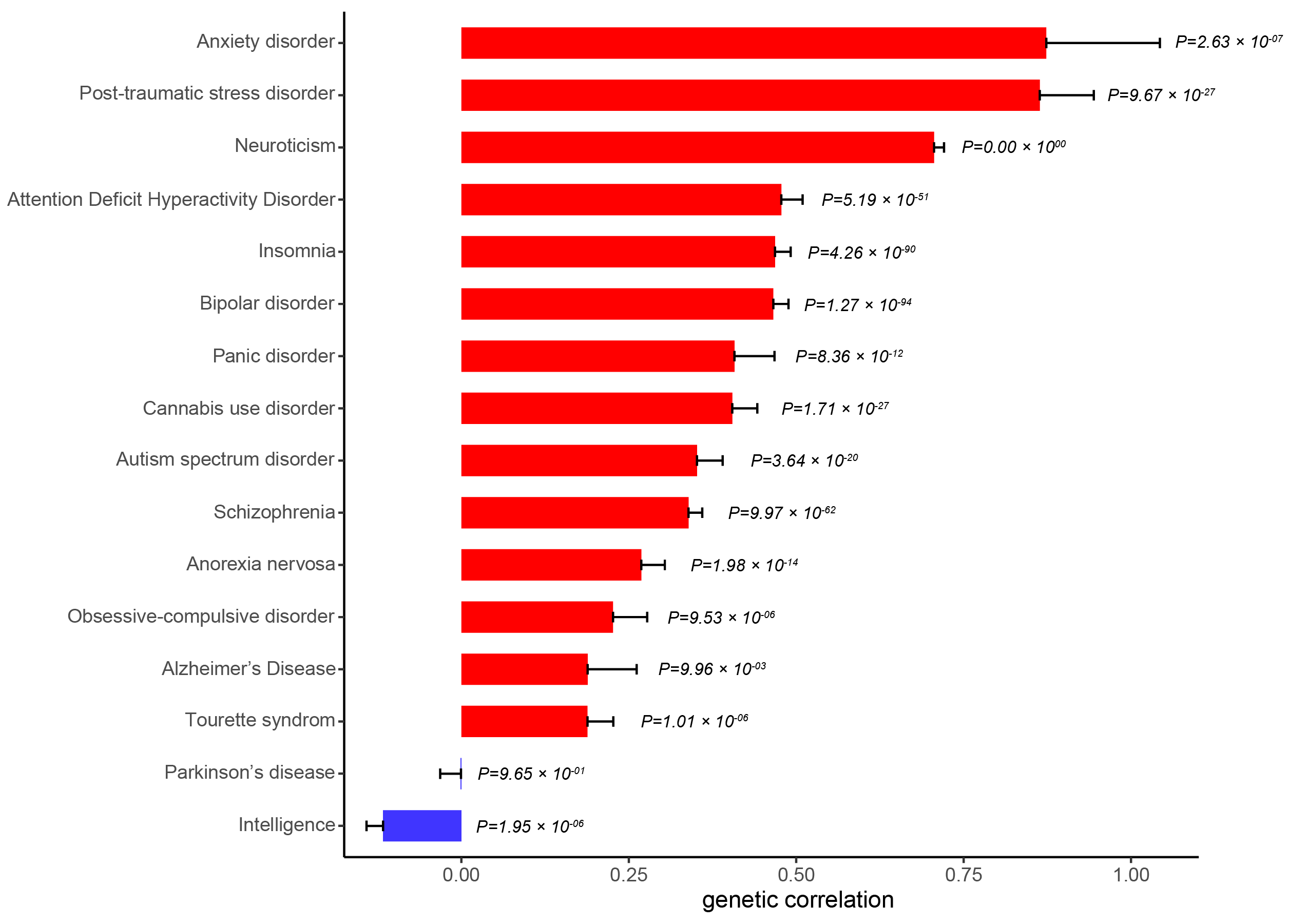

**Fig. S3. Genetic correlations between major depressive disorder and other brain disorders and intelligence.** Anxiety disorders show the most significant genetic correlation with depression.

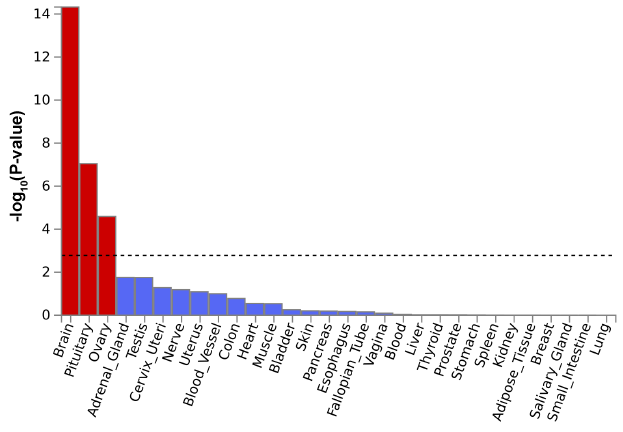

**Fig. S4. Heritability enrichment analysis in 30 general tissue types.**

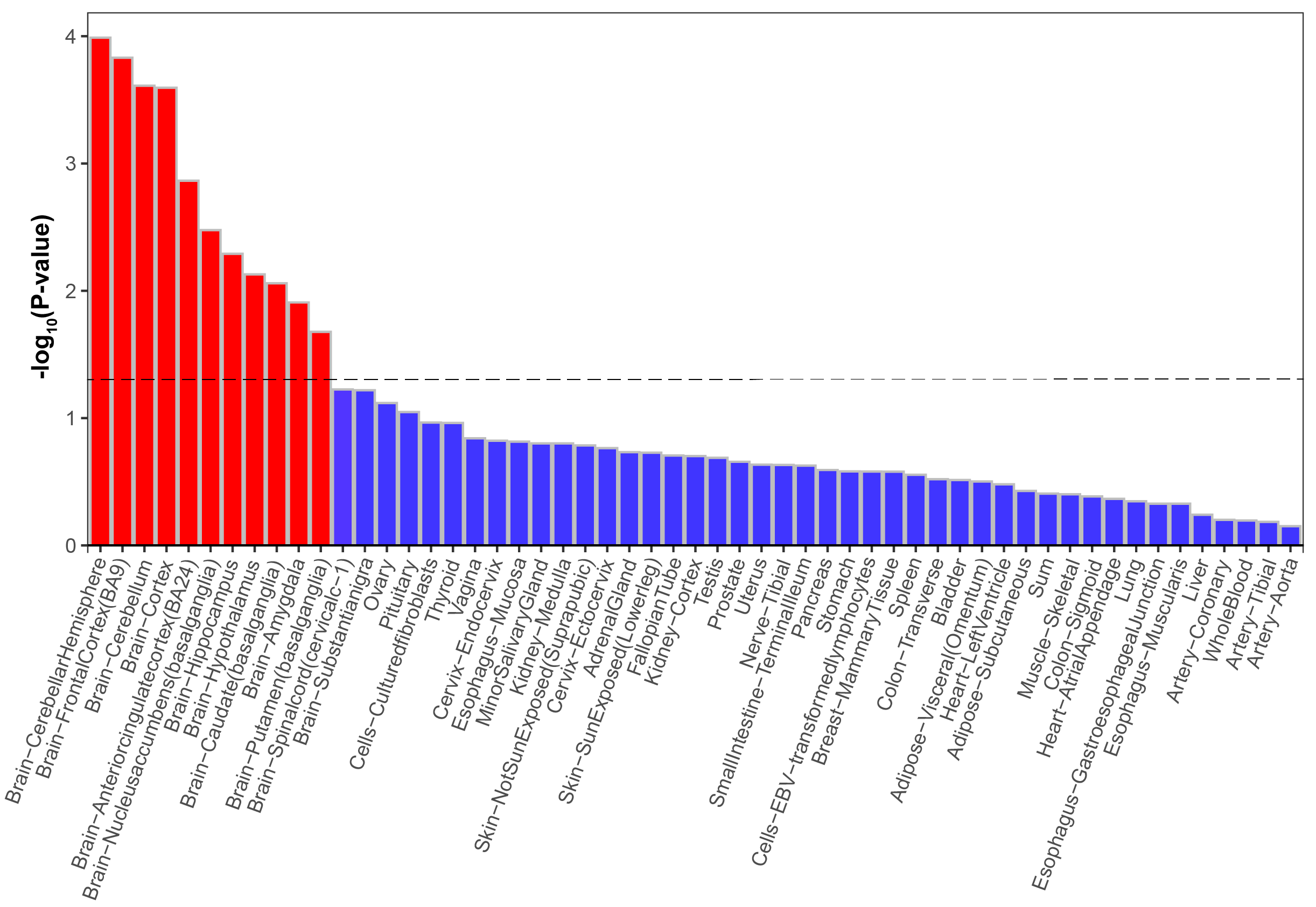

**Fig. S5. Heritability enrichment analysis in GTx V8 detailed tissue types.**

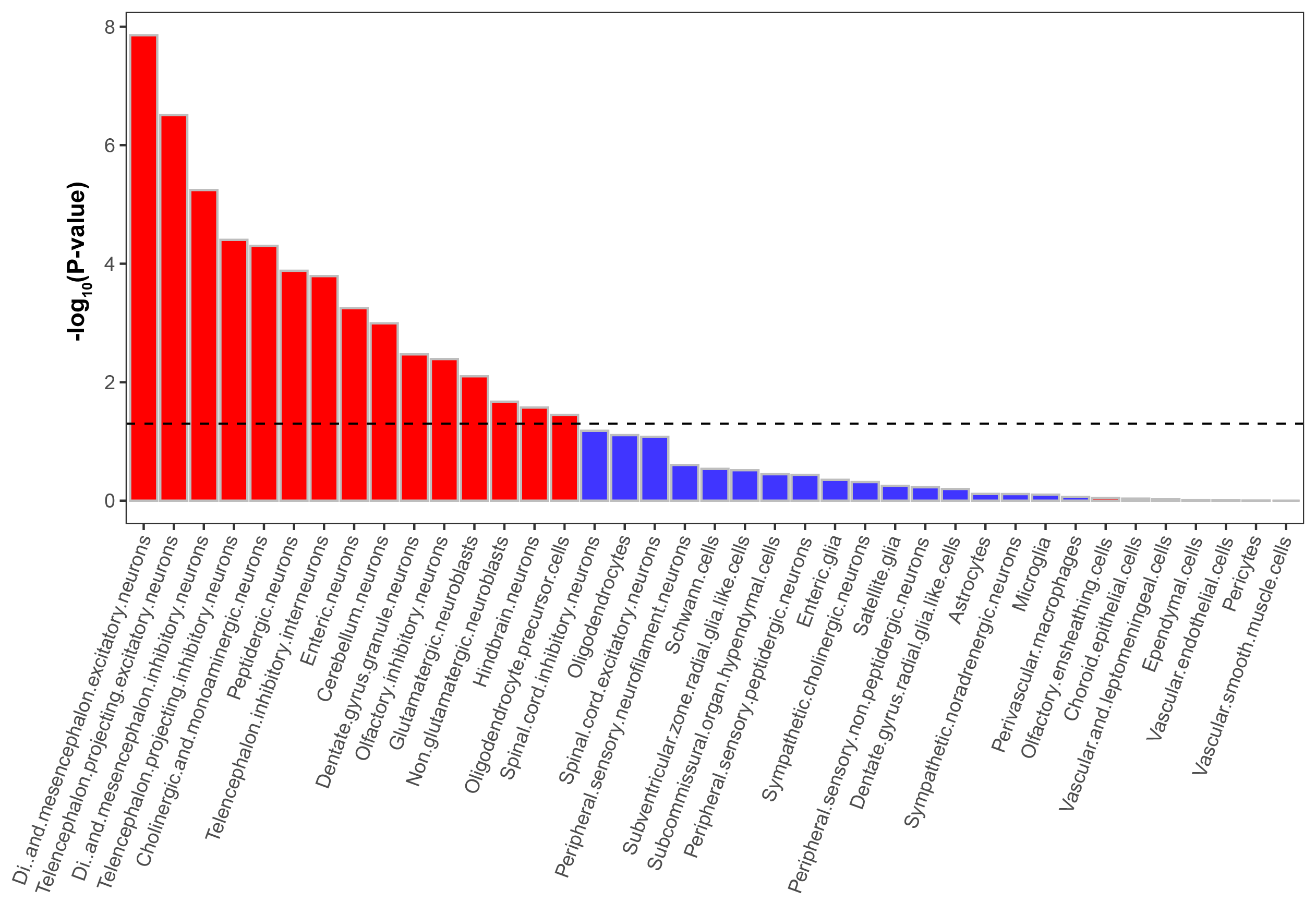

**Fig. S6. Heritability enrichment analysis in different cell type types.**

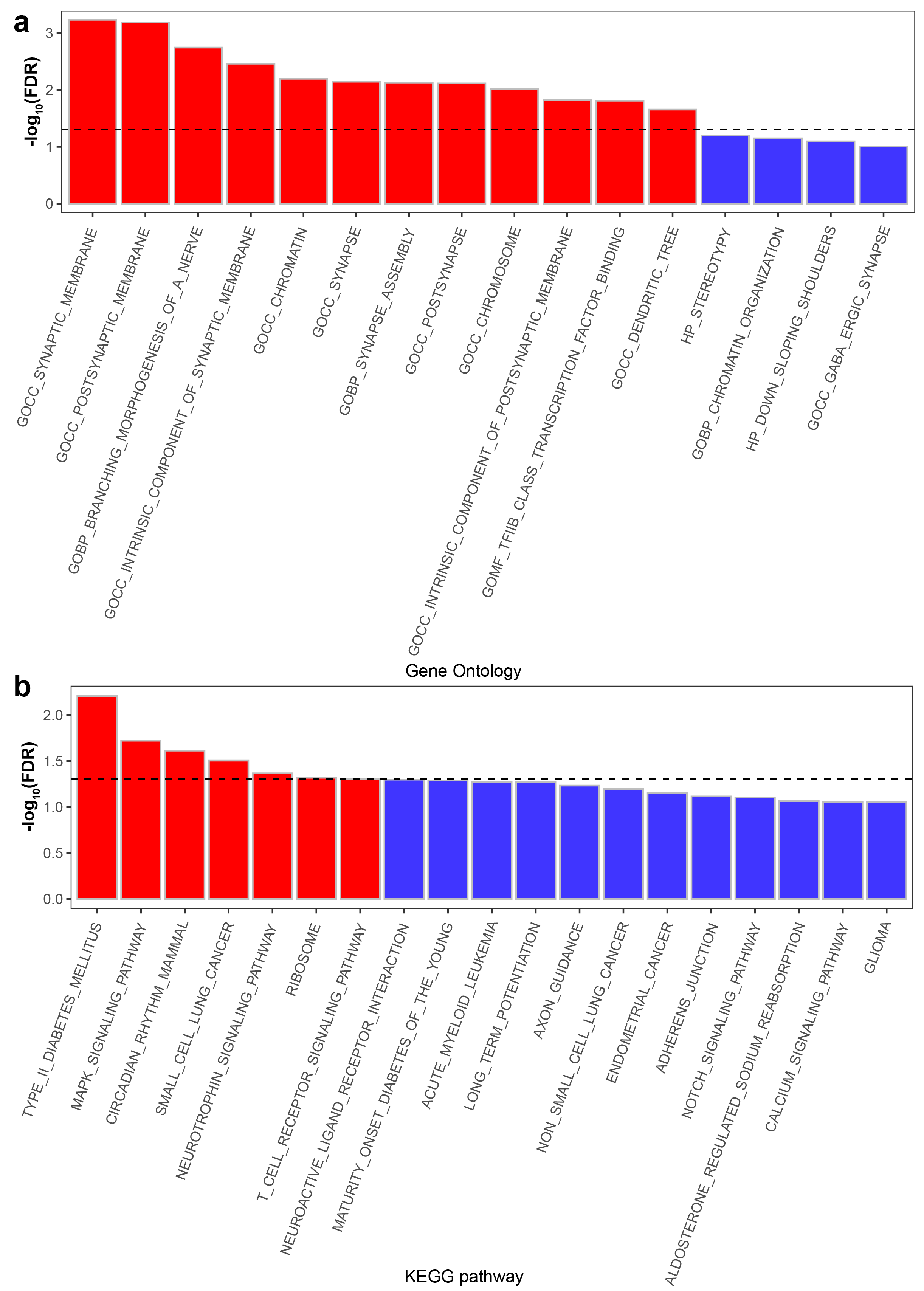

**Fig. S7. Heritability enrichment analysis of Gene Ontology (GO) and KEGG terms.**

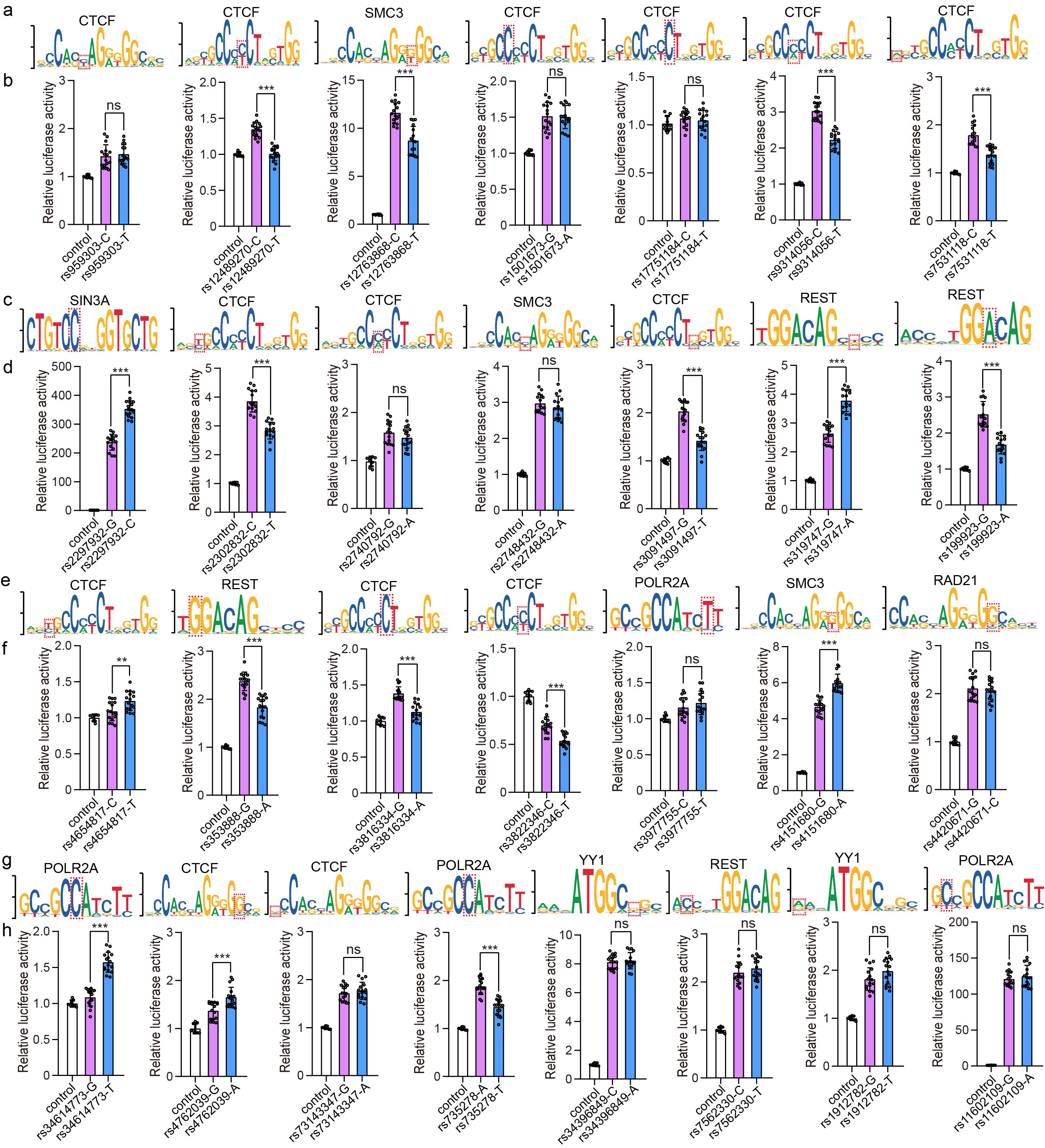

**Fig. S8 Reporter gene assays validated the regulatory effect of the identified TF binding-affecting SNPs.** Reporter gene assays validated the regulatory effect of the identified TF binding-disrupting SNPs. N = 8 for the control group, n = 16 per experimental group for SH-SY5Y cells. Two-tailed Student’s t test was used for statistical analyses. *P < 0.05, **P < 0.01, ***P < 0.001.

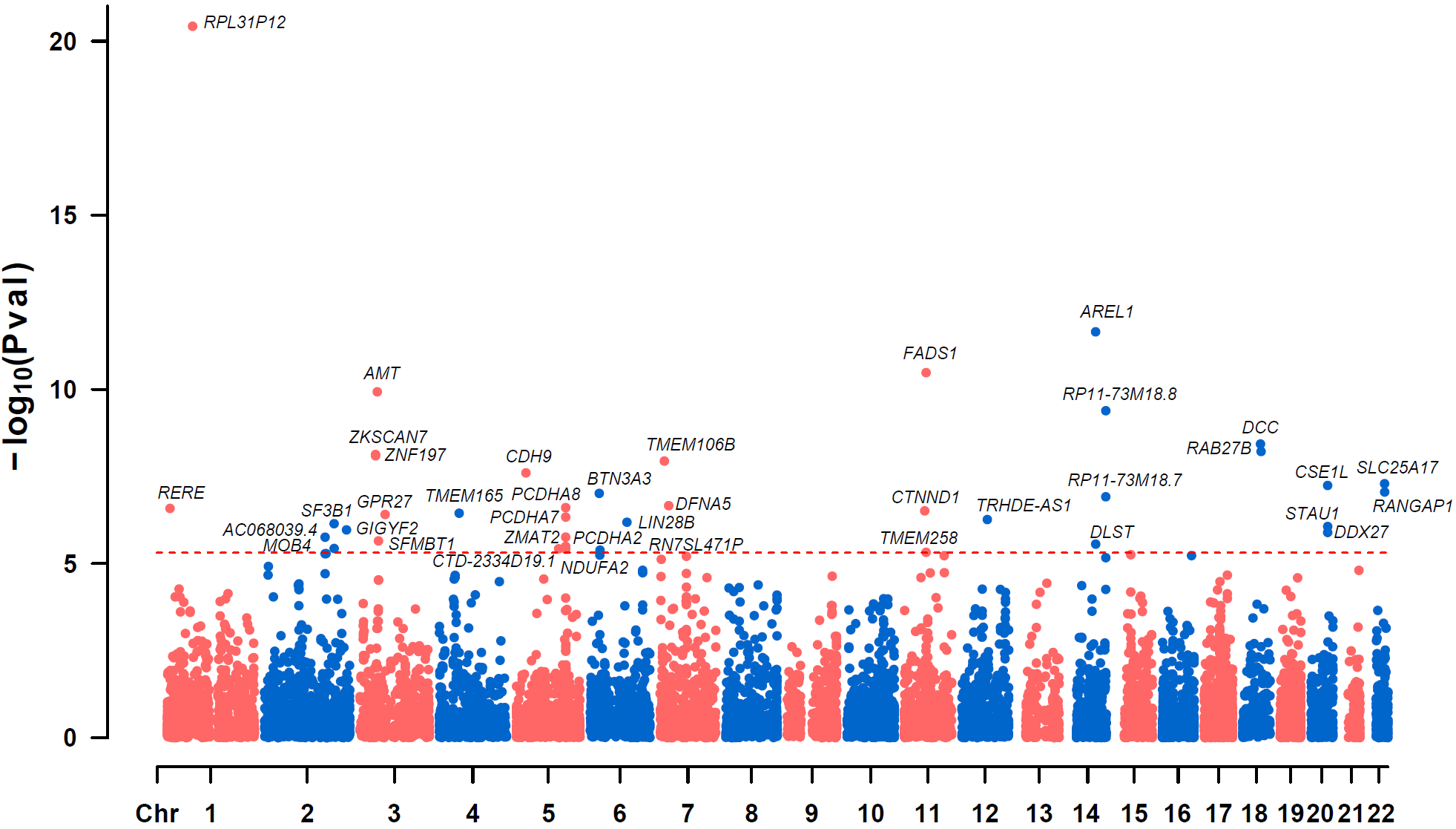

**Fig. S9. SMR analysis result using PsychENCODE cis-eQTL data.** The red line shows the Bonferroni-corrected significance threshold (0.05/10243, 4.88 × 10^-6^). Only MR-significant genes are shown.

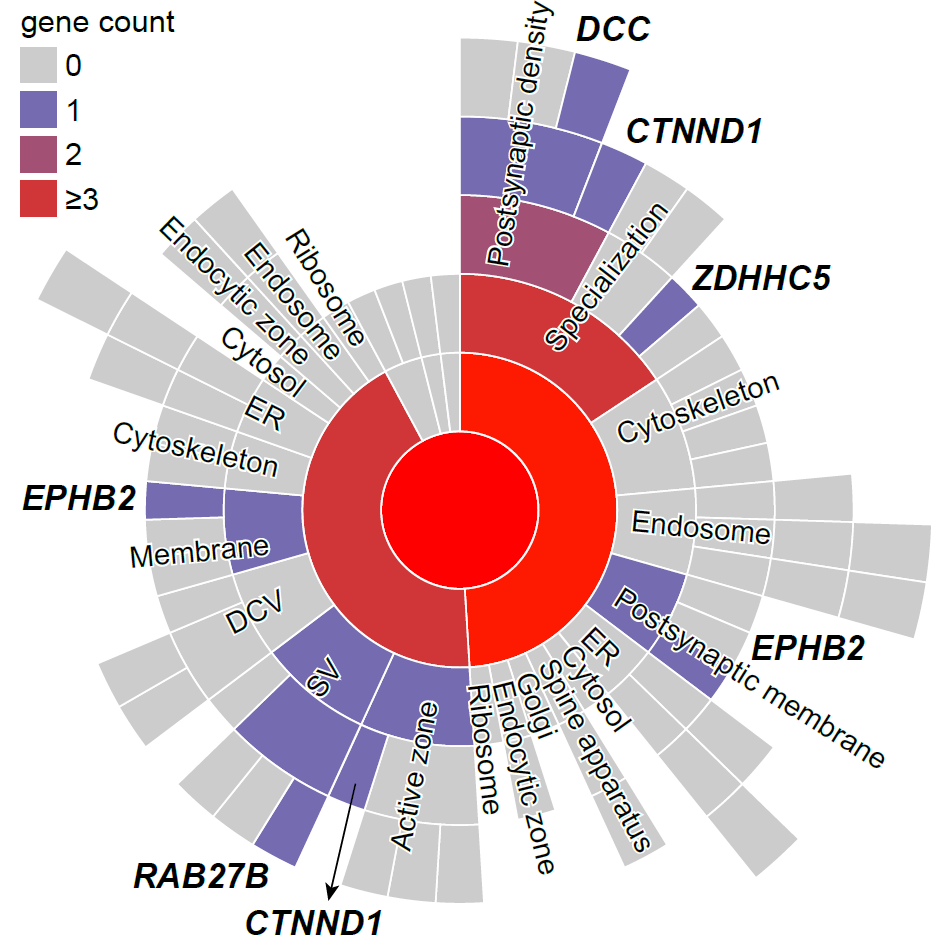

**Fig. S10. MDD risk genes in SynGO cellular components annotations.** The colors show the number of MDD risk genes annotated in SynGO. ER: endoplasmic reticulum; SV: synaptic vesicle; DCV: neuronal dense core vesicle.

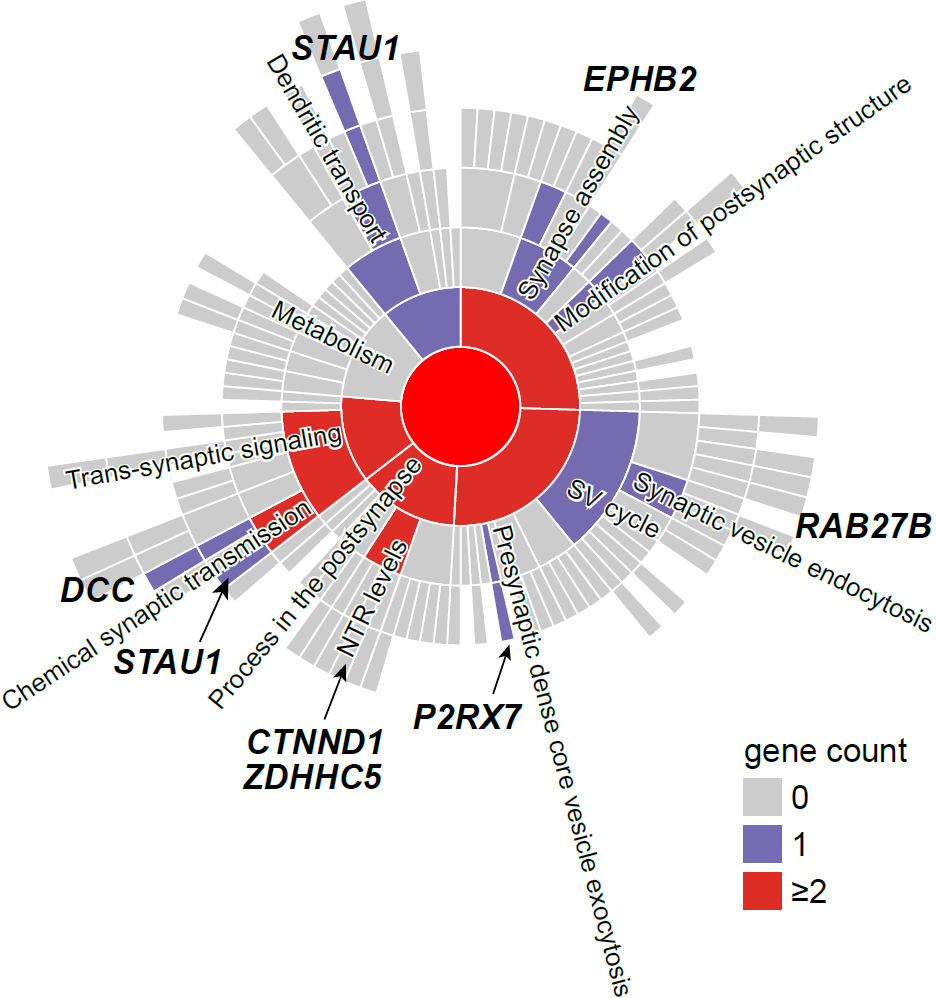

**Fig. S11. MDD risk genes in SynGO biological processes annotations.** The colors show the number of MDD risk genes annotated in SynGO. SV: synaptic vesicle; NTR: neurotransmitter receptor.
